# Antibiotic prescribing patterns by age and sex in England: why we need to take this variation into account to evaluate antibiotic stewardship and AMR selection

**DOI:** 10.1101/2024.09.10.24313389

**Authors:** Naomi R Waterlow, Tom Ashfield, Gwenan M Knight

## Abstract

**Objectives:** The drivers of antimicrobial resistance (AMR) likely vary substantially by different demographics. However, few complete open national detailed data exist on how antibiotic use (ABU) varies by both age and sex.

**Methods:** Here, prescriptions of antibiotics from General Practices in England for 2015-2023 disaggregated by 5-year age bands and sex were analysed at the national and Integrated Care Board (ICB) level. From a total of 249,578,795 prescriptions (across 9 years), 63% were given to women and the most prescribed were amoxicillin, nitrofurantoin and flucloxacillin sodium. Prescriptions per 100K population varied substantially across sex, age, geographical region, season, year, COVID-19 pandemic period and drug.

**Results:** Most antibiotics were prescribed more to women across most age bands (84% of antibiotics had more prescriptions to females across 50% of age bands). We show how this variation requires a more nuanced approach to comparing ABU across geographies and highlight that AWaRe targets are not met uniformly (young men have a higher proportion of “Watch” antibiotic prescriptions). We also show the impact on ABU of time-sensitive interruptions, including differential age-targeted influenza vaccination, COVID-19 restrictions and a shortage of amoxicillin combined with a Streptococcus A outbreak. Comparing to open access AMR data (MRSA in bloodstream infections) highlights the complexity of the link between ABU and AMR.

**Conclusions:** These detailed differences in ABU across England suggest that there should be large variation in AMR burden by age and sex, which now need to be quantified with detailed open access AMR data for a better intervention design.

## Introduction

The global public health priority that is antimicrobial resistance (AMR) is driven by a population-specific combination of antibiotic selection and transmission. There are many global targets that focus on the reduction of antibacterial use (ABU). One such target already adopted by the WHO 13^th^ General Programme of Work is to have at least 60% of total antibiotic consumption within the Access group of AWaRe antibiotics ^1–3^. More ambitiously, it has been proposed that 80% or more of oral antibiotic use could be Access ^2,4^. To achieve such targets without affecting patient care, it will be key to produce nuanced information about ABU across different subpopulations.

Most antibiotics are given outside of hospital settings ^5,6^ and previous studies have suggested that primary care antibiotic use (and hence exposure) varies substantially by age and by sex. A systematic review of primary care prescribing in 2016 suggested that women in high income settings receive a significantly higher number of prescriptions, particularly in the 16 to 54 years age band ^7^. This is supported by sub-national analyses^8–10^ from the UK, for example approximately twice as many antibiotic prescriptions given to women in 2013-2015, and 70% of all prescriptions after excluding antibiotics used to treat urinary tract infections (UTIs) ^8^. Antibiotic exposure rates are often “hockey-stick” shaped across age, with high levels in young children and an exponential increase with age ^7,11^. This is likely to reflect that different age/sex sub-groups present with different rates of infections^12^ and hence some level of appropriate prescription variation. For example women have higher rates of UTIs ^13^.

Antibiotic use also varies geographically at national and sub-national levels ^14–16^. Identifying outliers once patient composition is accounted for, is vital for improving antibiotic stewardship^17,18^. To do this, various measures have been proposed such as the English STAR-PU ^19^ which uses different prescribing weights in different ages and sexes. Issues with this metric include the lack of accounting of co-morbidities ^18^ and the grouping of all antibiotics together masking variation in antibiotic use due to patient or GP-related factors ^20^, although variation in prescribing may be decreasing over time ^21^.

To reduce ABU many age-specific interventions could be considered, for example vaccination programmes are postulated to reduce both unnecessary and necessary antibiotic use ^22,23^. Evidence that such a reduction in antibiotic exposure leads to reduced resistance is mixed^24^ , but age- and sex-disaggregated ABU could be used to reveal where there is a link between vaccination and ABU, as well as ABU and resistance rates. However, few ecological open access ABU or AMR data exist disaggregated by both age and sex. In one of the only detailed age and sex analyses, we have previously shown that AMR prevalence in bloodstream infections in Europe varies substantially by age, sex and bacteria-antibiotic combination ^25^.

Here we use 9 years of open access national data on antibiotic prescriptions from primary care in England available at an unprecedented level of both age and sex disaggregation to reveal the variation in ABU. We test assumptions about antibiotic weightings used for identifying prescribing outliers, conduct a nuanced analysis of AWaRe targets, generate estimates of the impact on ABU of age-based influenza vaccination, and explore the link between ABU and AMR. The patterns we find show the importance of such nuanced data and emphasise the need for more disaggregated AMR data.

## Materials & Methods

The number of primary care antibiotic prescriptions by 23 age bands and 3 genders in England & Wales was received through two freedom of information requests (FOI-01671, FOI-01975) ^26^ to the NHS Business Service Authority (NHSBSA) Information Services Data Warehouse containing information from the NHSBSA Prescription Services. The first FOI contains data at General Practice (GP) and Integrated Care Board (ICB) level for England and some of Wales. The second is total national English data only.

This data covers the 9 year period of April 2015 to December 2023 inclusive, by month, and includes all medicines from the British National Formulary (BNF) section 5.1: Antibacterial drugs ^27^. Datapoints relating to fewer than 5 items prescribed were redacted by NHSBSA for privacy reasons. The 23 age bands were “0-1”, “2-5” and then 5yr age bands up to “105+”. The three gender classes included were “Male”, “Female” and “Indeterminate” (where information does not confirm either Male or Female). We used number of items prescribed in our analysis, so as to avoid double-counting of patients ^26^.

Of total prescriptions in the dataset, 3.3% were prescribed to a patient of unknown gender or age band. We excluded these prescriptions from our analysis, although note that the frequency varied with time, with fewer unknowns over time (Appendix 1 section 2). We also excluded prescriptions given to individuals of “Indeterminate” gender, due to the small numbers (6247 Items, 0.0024%) and the antibiotics for which there were less than an average of 10 prescriptions per year (22 antibiotics, Appendix 1 section 3). For our main analysis, we assumed all redacted numbers of items had a value 1 (from a possible 1,2,3 or 4), and run sensitivity analyses on this number (Appendix 1 Section 7).

Population sizes by age were obtained from the Office for National Statistics (ONS) and mid-year estimates were used for each calendar year^28,29^ (Appendix 1 Section 4). We only included prescriptions for England in the analysis, and only those for 2023 were used for sub-national analyses, due to changes in the definitions of the ICBs. Data from NHSBSA provides “Gender”, but most other sources use “Sex”. We have assumed that we can match these two in this analysis.

We calculated an updated STAR-PU weighting of antibiotic prescriptions per age band and sex, with smaller age bands ^19^. The updated comparison metric (UCM) weightings were calculated using females aged 66-70 as baseline (a subset of the STAR-PU 65-74 female baseline). Our UCM weightings were then applied to the ICBs in 2023, using the same STAR-PU principles: the new UCM weightings were multiplied by the relevant populations in each ICB, and the total prescriptions per UCM-population were used to compare ICBs. In addition, drug family-based weightings were calculated using the same methodology (“UCM-family”), but prescriptions from each drug family were used. For children aged up to 16 (the only age groups where there were sometimes no prescriptions) a value or 0.01 was used if there were no data in that age band, and limits were set of 0.01 to 100.

Drugs were classified into the AWaRe categories using a World Health Organisation reference table ^30^. Where drug names were not identical, these were manually matched (Appendix 1 section 1).

Age bands covered by influenza vaccination were extracted from the annual UKHSA reports ^31^, and these were compared to prescriptions of antibiotics used in respiratory tract infections (RTIs)(defined as all drugs in the BNF categories “cephalosporins and other beta-lactams” and “penicillins”), excluding amoxicillin. Amoxicillin was excluded due to the very large number of prescriptions, and it’s use in many varieties of infection^32^. The influenza season was defined as 1st of October to 30th March inclusive. The number of prescriptions given to patients in age bands that matched closely to the vaccination groups was calculated where possible (0-1, 2-5, 5-10, 11-15, 50-65, 65+).

We extracted the only publicly available data on AMR infection split by age and sex. This was for *Staphylococcus aureus* bloodstream infection data from UKHSA ^33^. The proportion of these BSIs due to *S. aureus* that were methicillin-resistant (MRSA) were compared to prescriptions of antibiotics in the BNF categories “cephalosporins and other beta-lactams” and “penicillins”.

Data collection and transmission rates were altered during the years of SARS-CoV-2 circulation and lockdowns, which we classified to be from March 2020 to June 2022 inclusive for England.

All code is available on github (https://github.com/NaomiWaterlow/prescription_foi) and was conducted in R ^34^.

### Sensitivity analysis

The main issue to explore in the data was the uncertainty in the redacted small numbers (“*”) when the number of prescriptions was between 1-4. In sensitivity analysis we explored changing our default value of 1 to 4 for (a) everyone, (b) males only and (c) children only (< 20yos).

## Results

### Prescription data

Over the 9-year period April 2015 to December 2023, 249,578,795 prescriptions of antibiotics were given in England. For the complete years (2016-2023), the average was 0.48 (range of 0.43, 0.52) prescriptions per person per year. The most frequently prescribed drugs were amoxicillin, nitrofurantoin and Flucloxacillin sodium(Table 1). 63% (63%, 64%) of drugs were prescribed to females, with 0.60 (0.54, 0.65) per person per year versus 0.36 (0.31, 0.40) in men. 13% (10%, 15%) of prescriptions were given to those <16yo, and 35% (33%, 37%) to those over 65yo. The highest overall prescription rate in both sexes was in the 86yo+ band (Appendix 1 section 5). COVID-19 mostly affected prescribing in those aged < 16yos (Appendix 1 section 5), mostly returning to pre-COVID-19 levels in the Spring of 2021.

**Table 1:**
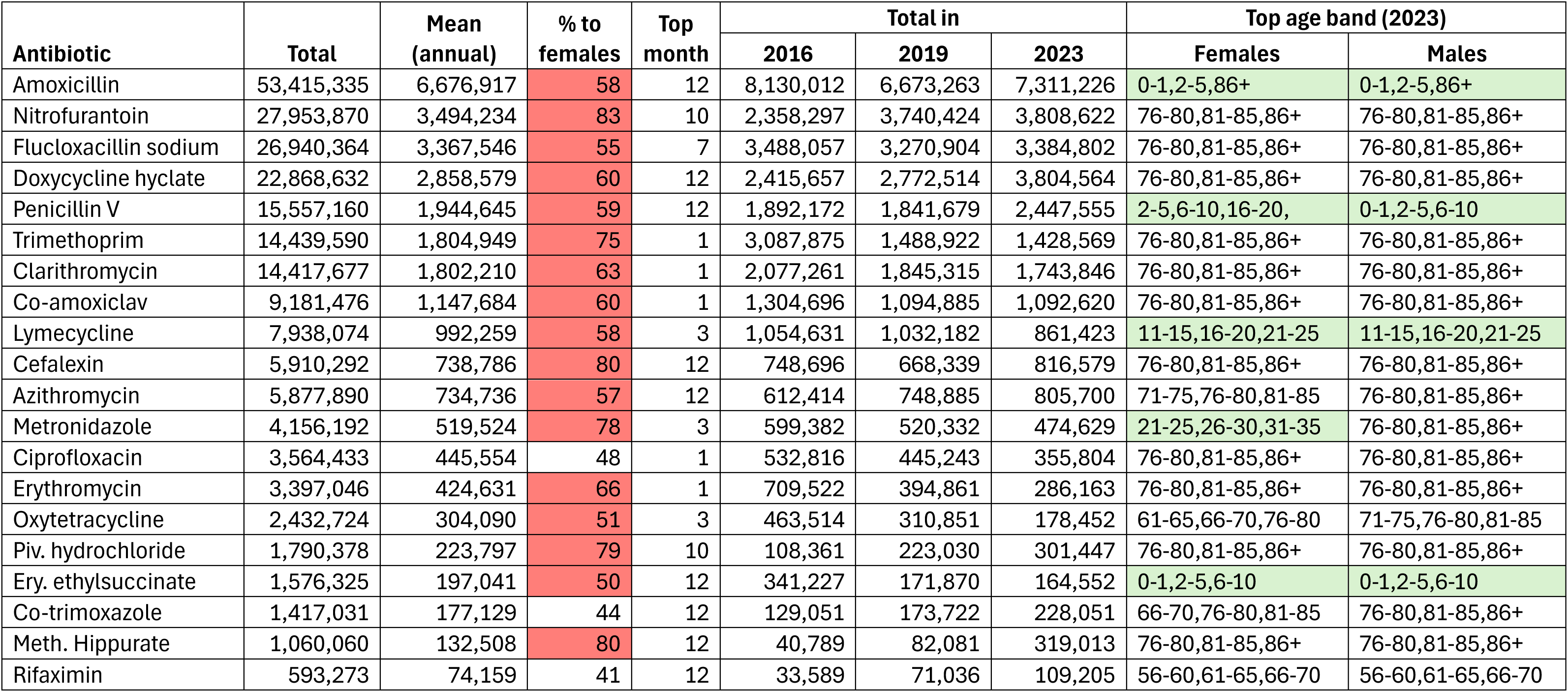
Summary data on number of prescriptions for the 20 most prescribed antibiotics (see *Appendix 1 section 6* for all antibiotics) across the complete years of the data (2016-2023) unless otherwise indicated. Red shading indicates more prescriptions are to females and “Top” month is the month with the most prescriptions of this antibiotic (1 = January), when summing across the whole dataset. “Top age band (2023)” is the age band in 2023 with the highest prescription rate per 100k. Light green shading indicates relatively more prescriptions to younger individuals vs top age bands all being 55yo or more (Ery. is Erythromycin. Co-amoxiclav is amoxicillin and clavulanic acid, Co-trimoxazole is trimethoprim and sulfamethoxazole. Piv. = Pivmecillinam. Meth. = Methenamine.)

The top 20 prescribed antibiotics, totalling 98% of prescriptions, (Table 1) had annual prescription totals varying from 74,159 to over 6.5 million prescriptions, with most being prescribed more commonly to females (17/20) and most often in December (9/20) or January (5/20). Those prescribed more to men were ciprofloxacin, co-trimoxazole and rifamixin. Just over half of the top 20 prescribed antibiotics had fewer prescriptions in 2023 than 2016 (11/20), with 61% of antibiotic having at least a 10% reduction in prescriptions between 2019 and 2023, although some had sex-based time variation (e.g. ofloxacin, Fig 1B). Most of the top 20 antibiotics (15/20) had the highest rates in older ages (>55yos) in 2023.

**Figure 1:**
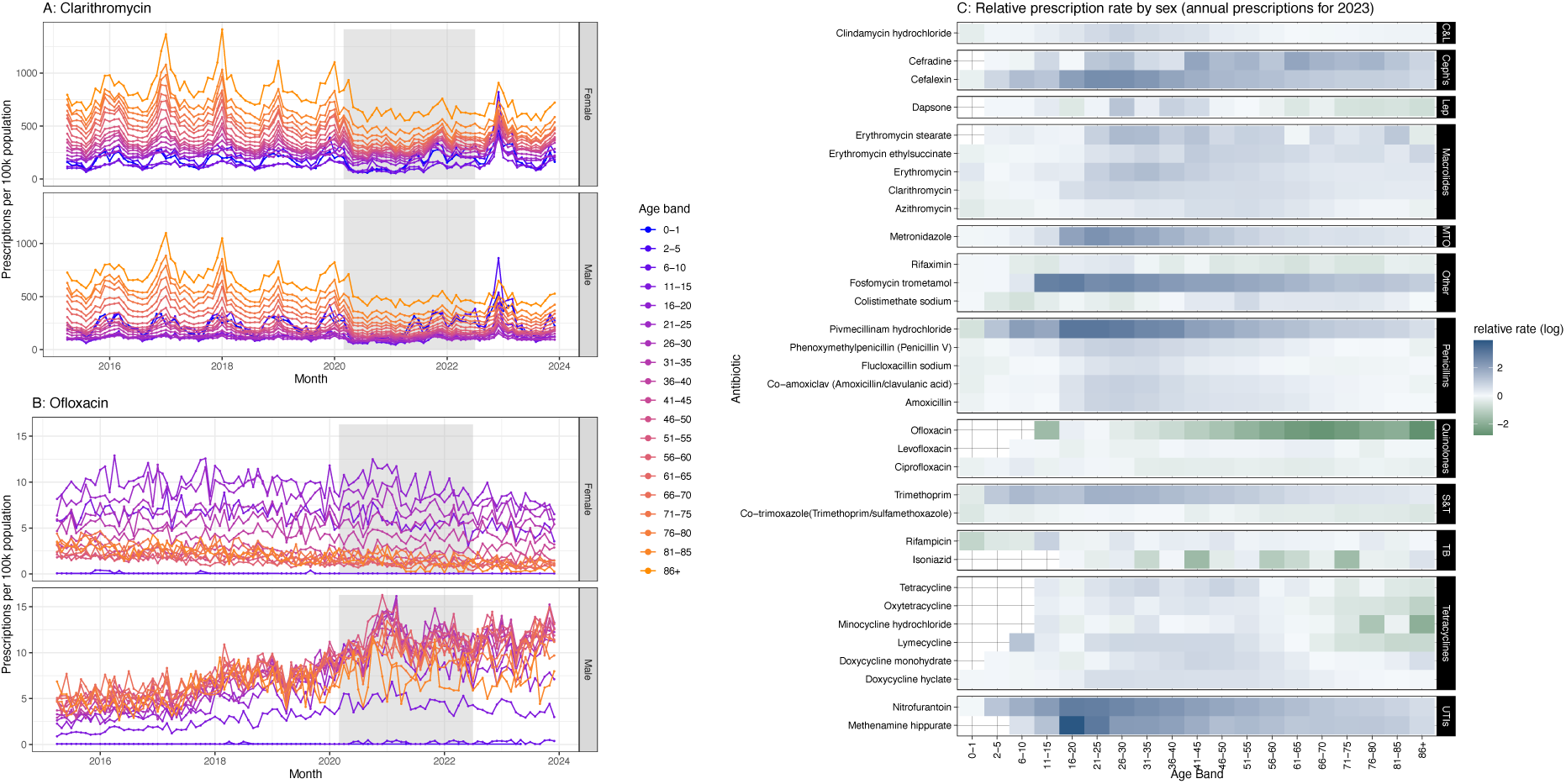
Variation in antibiotic prescribing by age and sex with examples and overall comparison A/B). Example prescription rate per 100’000 population for clarithromycin and ofloxacin respectively. Colours indicate age bands, facets indicate sex. Grey shading indicates years of COVID-19 interventions. See *Appendix 2* for all antibiotics. C) Relative prescription rate by sex for prescribed medicines in 2023. Colours represents the log of the ratio, with blue indicating higher prescriptions in women, and green indicating higher prescriptions in men. Drugs are ordered by BNF classification, where the acronyms are: Clindamycin and lincomycin = “C&L”, Cephalosporins and other beta-lactams = “Ceph’s”, Antileprotic drugs = “Lep”, Metronidazole, tinidazole and ornidazole = “MTO”, Sulphonamides and trimethoprim= “S&T”, Antituberculosis drugs = “TB”, Urinary-tract infections = “UTIs”.

Twenty-eight antibiotics (38%) show an annual seasonal pattern (Fig 1A, Appendix 2) in prescriptions (pre-COVID-19), defined as having at least two months where prescriptions were 20% higher than the average number of prescriptions, averaged over years. These seasonal antibiotics, all had a decrease in prescriptions during the COVID-19-period, followed by a spike in late 2022 (Appendix 2) correlating with a Streptococcus A outbreak and amoxicillin shortage^35^.

For most age bands higher levels of prescription in females compared to males (Fig 1C), were observed (84% / 54% / 24% of antibiotics had more prescriptions to females in at least 50% / 75% / 90% of age bands respectively). Notable exceptions with higher prescriptions to males are quinolones, isoniazid and minocycline hydrochloride. For some ages the sex that receives the most prescriptions changes with age (e.g. colistimethate sodium, azithromycin and rifampicin have higher prescriptions in males in children, higher prescription rates in females in adults (Fig 1C)).

#### Antibiotic usage metric

We created an updated comparison metric (UCM), with smaller age bands than STAR-PU (Figure 2B). This resulted in more extreme values across age bands, ranging from 0.19 to 2 in our UCM, compared to a range of 0.2 – 1.3 in the 2013 STAR-PU with particularly higher values in older males (Figure 2A vs B). Values by BNF drug value varied considerably by age and sex (“UCM-family”, Appendix 1 section 7).

**Figure 2:**
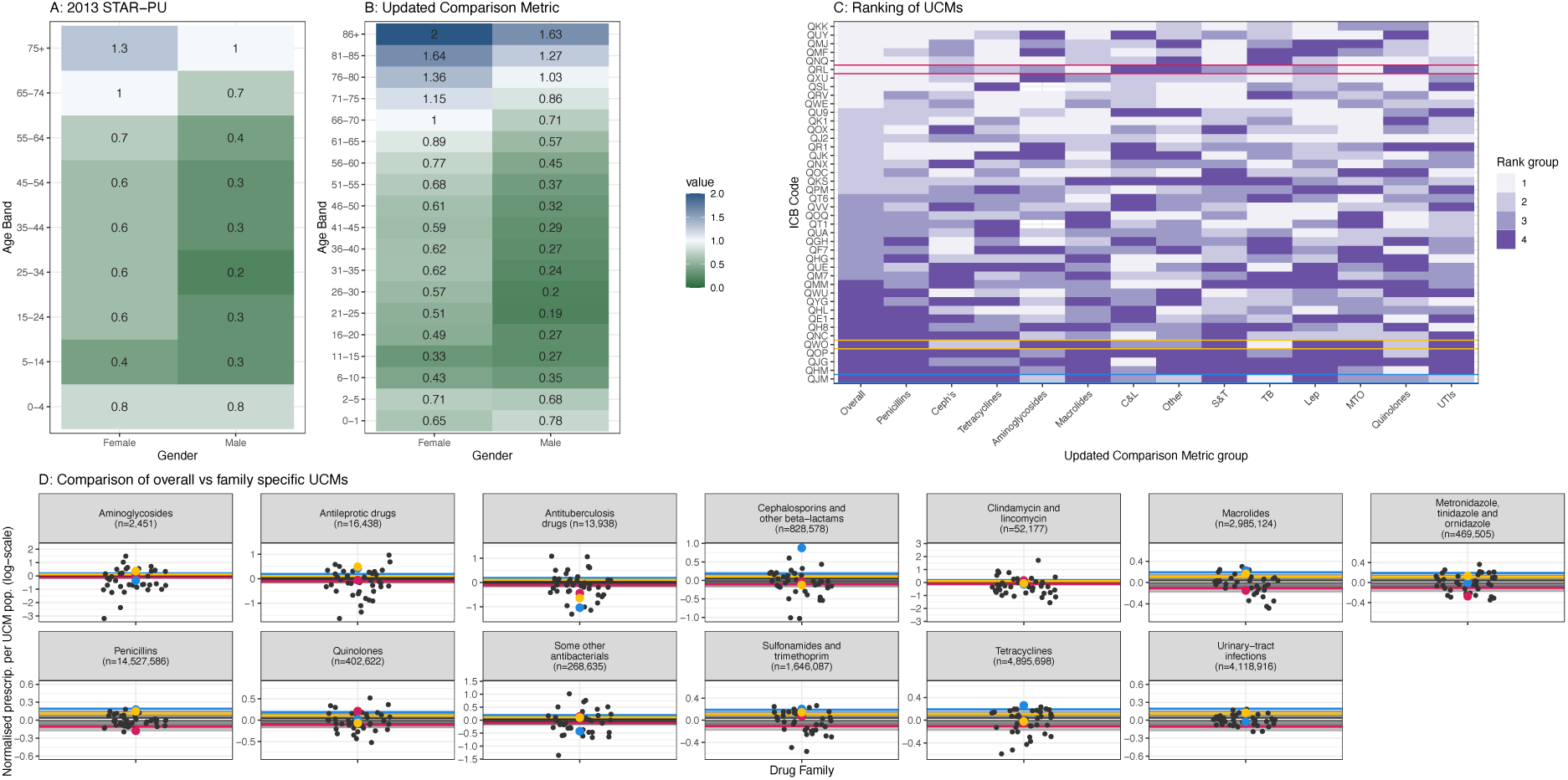
A)2013 STAR-PU values using England data. B) Updated Comparison Metric (UCM) using 2023 English data and more disaggregated age bands. Blue = high value, green = low value, centred around the baseline: females, 66-70 (white). C) Ranking of UCMs across English ICBs, grouped by colour into quarters: rank 1-10 (1), 11-20 (2), 21-30 (3) and 30-42 (4). The “Overall” column shows the ranking of ICBs using our UCM, and the other columns by UCM-family. Acronyms: Clindamycin and lincomycin = “C&L”, Cephalosporins and other beta-lactams = “Ceph’s”, Antileprotic drugs = “Lep”, Metronidazole, tinidazole and ornidazole = “MTO”, Sulphonamides and trimethoprim= “S&T”, Antituberculosis drugs = “TB”, Urinary-tract infections = “UTIs”. D) Normalised prescriptions per English ICB and antibiotic (facet) for overall UCM (lines, same on each facet) and UCM-families (dots). In C) and D) the same three ICBs are highlighted with colour (blue, yellow and pink). n indicates the total number of prescriptions across all ICBs.

The new-UCM ranking of ICBs (first column Fig 2C, lines in Fig 2D), changed substantially when applying the UCM-family (Fig 2C, points in Figure 2D). For example, ICB QRL ranks as a high prescriber with the new-UCM (6^th^ overall, Fig 2C, red in Fig2C&D) but a low prescriber of quinolones (38^th^ overall, Appendix 1 section 9). The range in UCM-family ranking decreased with more prescriptions (Fig 2D).

#### Antibiotic consumption targets

The WHO 60% “Access” prescribing target has been reached in England: 84% of primary care prescriptions were “Access” in 2023. However, the more ambitious 80% level is not being met in males ages 11-20, largely driven by a spike in prescriptions for lymecycline (Figure 3B), commonly prescribed for Acne ^36^. The main Reserve drug prescribed by GPs in 2023 was colisitimethate sodium, which must be given by injection or as a nebuliser as it is not given orally ^37^, likely under the direction of tertiary care, particularly for Cystic Fibrosis patients ^38,39^. The percentage of antibiotics that were “Access” varied across ICBs by between 5.3% (females, aged 6-10) to 13.4% (males, aged 16-20) (Appendix 1 Section 10).

**Figure 3:**
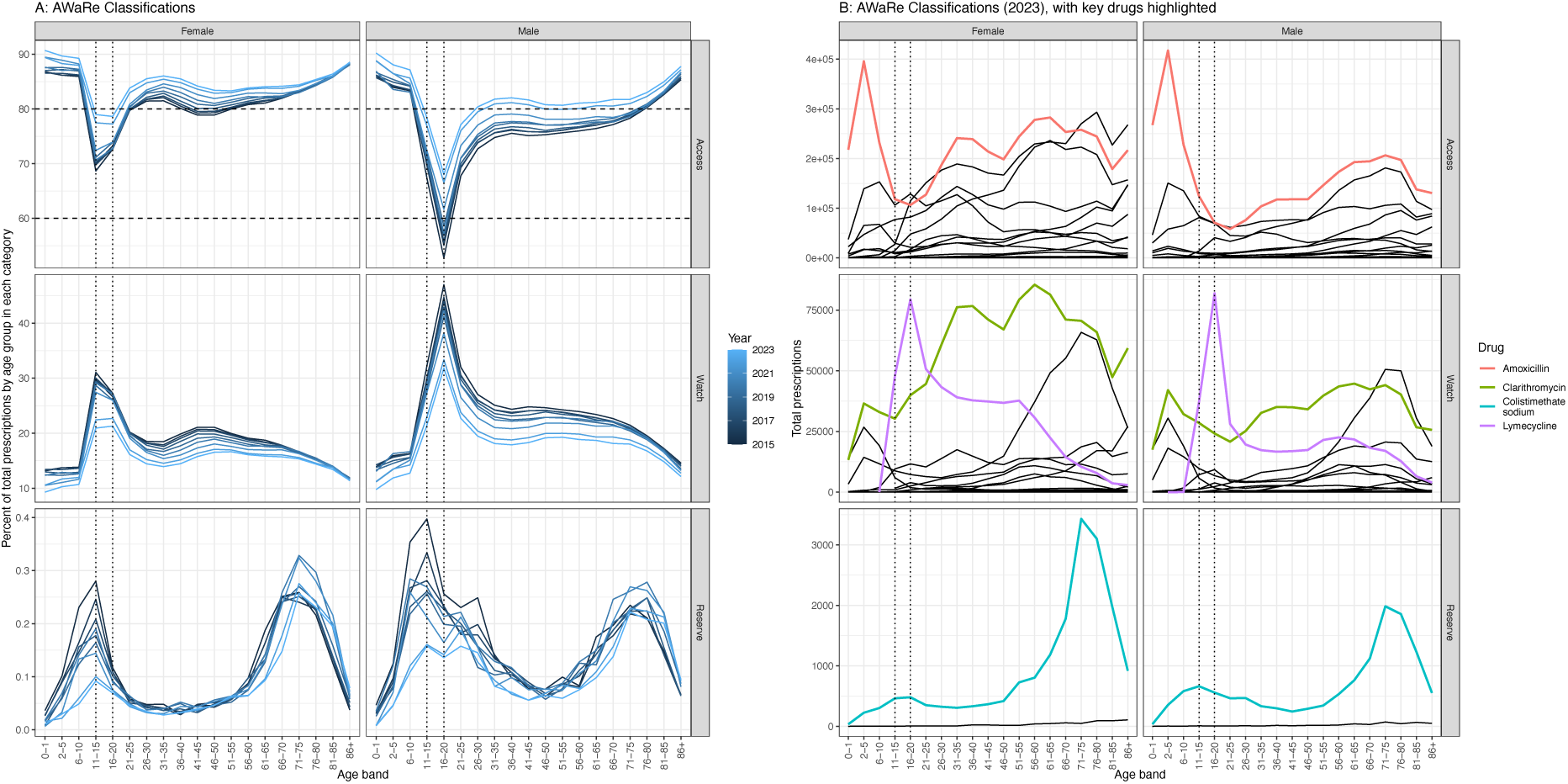
A) Percentage of total prescriptions in each of the AWaRe categories (row), by age band, sex (column) and year (colour). Dashed horizontal lines represent targets of 60% and 80% in the Access category, and dotted vertical lines are included for comparison across graphs. Note the dijerence in y-axis ranges. B) Total prescriptions in 2023 by AWaRe categories (row), age band and sex (columns). Key contributing drugs are highlighted by colour and vertical dotted lines are included for comparison across graphs.

#### Influenza vaccination

Comparing the relative rate of prescriptions commonly given for (suspected) RTIs before and after influenza vaccination programmes (Figure 4A) shows no clear impact. For example, compared to ages 15-50, RTI relative prescription rates declined by just under 5% in age band 6-10 following introduction of vaccination in 7-8yos (Figure 4A&B).

**Figure 4:**
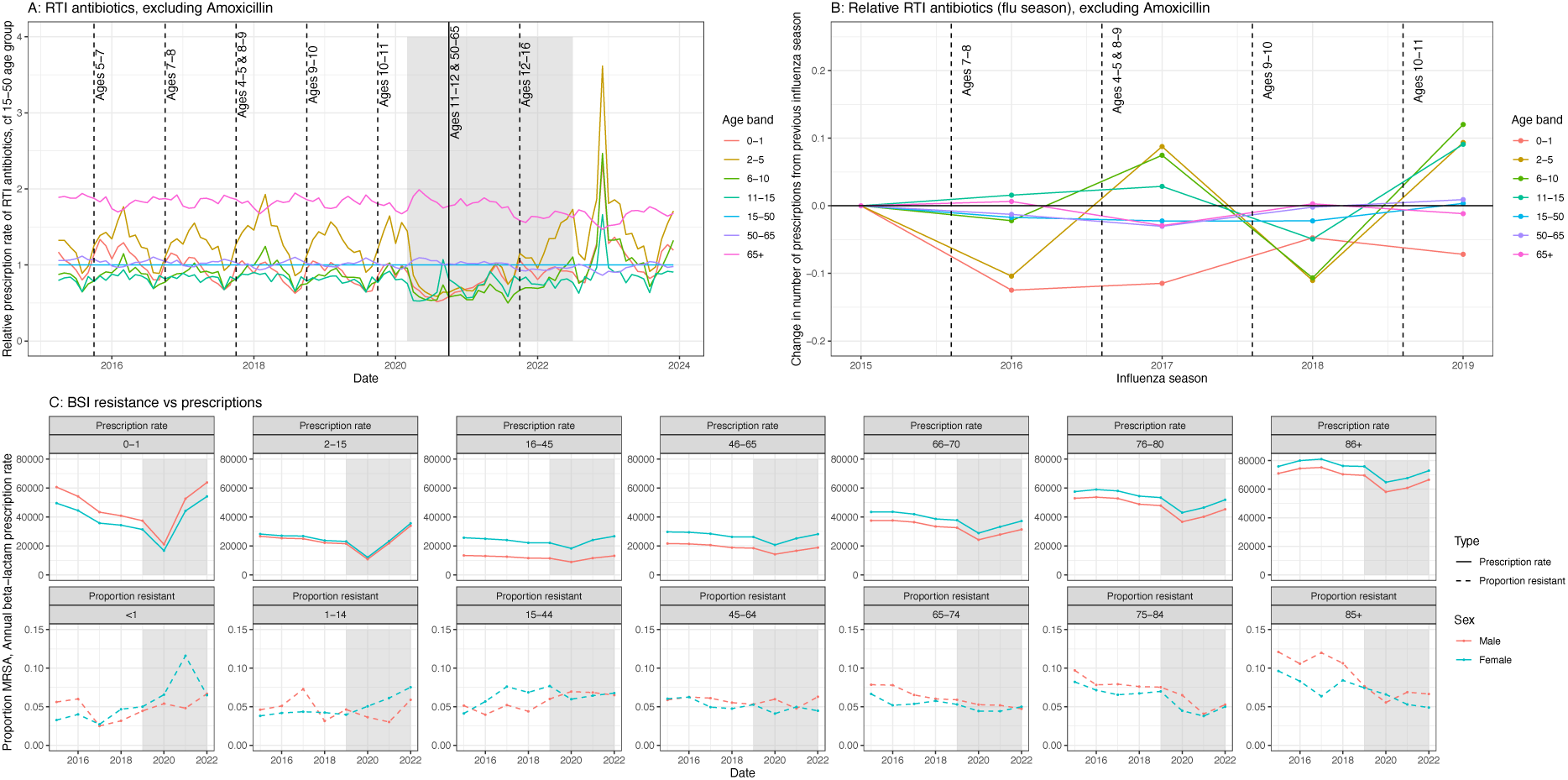
A) Age band specific prescription rates of RTI antibiotics (penicillins, cephalosporins and other beta-lactams) relative to the rate in age band 15-50. Amoxicillin was excluded (see methods). The influenza season was defined as 1^st^ of October to 30^th^ March inclusive. Vertical lines indicate the year the introduction of vaccination in the labelled age band, and the COVID-19 years are shaded grey. B) Proportional change in the number of RTI prescriptions (excluding amoxicillin) from the influenza season of the previous year, by age band. Vertical lines indicate the introduction of vaccinations in the named age bands C) Prescription rate of beta-lactam antibiotics (top row) and proportion of BSI due to resistant S. aureus (over all S. aureus BSI) for similar age bands (columns).

However, in this same period the prescriptions in age bands 0-1 decreased by over 10%. There was an increase in relative prescriptions in all ages except older adults when vaccination was introduced in 4-5yos & 8-9yos. Vaccination of 50-65yos would likely have the largest impact (more people), but is hard to interpret, as this occurred during the COVID-19 years. No decrease in prescriptions in this age band compared to the other age bands was identified (Fig 4A).

#### Impact on antimicrobial resistance (AMR)

There was a lack of a clear link between prescribing and resistance rates by age and sex for *S. aureus* (Figure 4C). All age bands except ages 0-1 see higher prescriptions of beta-lactam antibiotics (those to which MRSA are resistant) in females, yet the proportion of BSIs due to MRSA is higher in males 45yo and over, and varies over time in 1-44yo. As opposed to prescriptions, a higher proportion of BSIs were due to MRSA in females in age band <1 over time.

#### Sensitivity analyses

The majority of age bands across drugs show higher levels of prescription in females compared to males in all sensitivity analysis. When the hidden low prescription numbers (*s) were all set to the maximum possible value (4 instead of 1, equating to 0.13% instead of 0.033% of all prescriptions), 85% / 55% / 27% of antibiotics had more prescriptions to females in at least 50% / 75% / 90% of age bands respectively. The sensitivity analysis with the higher value (4) in males only or children only had highly similar levels of 78% / 55% /18% and 85% / 55% / 24% respectively (Appendix 1 section 7).

## Discussion

We report here, for the first time, national data on all antibiotic prescriptions in primary care in England disaggregated by both 5yr age bands and sex. This unprecedented level of detail highlights huge variation and will be vital in bringing a more nuanced approach to AMR control.

Whilst most antibiotics (15/20 of most prescribed antibiotics) had higher prescription rates in older ages (55yo+), a substantial proportion had higher prescription rates in children or younger adults. Matching previous reporting ^7–10^, women were prescribed more antibiotics, but this pattern was not uniform across age bands and the patterns seen would have been hidden if using only three age groupings. Higher prescribing in women is likely linked to both infection syndrome variation and to behavioural differences in consultation rates ^8,40–43^. We saw clear syndrome-linked prescribing: higher levels of UTI prescribing (nitrofurantoin and trimethoprim) in women and higher levels of prostate or gonorrhoea infection linked prescribing (ofloxacin) in men. Quantifying this age and sex variation provides a baseline for intervention targeting and may help assess prescription appropriateness.

Most antibiotics (∼60%) did not have a seasonal pattern of exposure. Those that did had a lower prescribing rate during COVID-19, likely reflecting that antibiotics prescribed seasonally are given for infections that are dependent on seasonal changes in transmission (schools / time indoors), or linked to other infections (e.g. respiratory viruses ^44,45^). Supporting this theory is the rapid increase in prescribing in late 2022 which coincided with a *Streptococcus A* outbreak and a wide increase in prevalence of respiratory virus infections linked to “re-opening” ^46^. How this could drive seasonality in AMR needs untangling. Other studies have found a similar decrease in prescribing during COVID-19 in primary care in sub-national data in the UK ^47 48^, mostly driven by prescribing in children.

These data allow for more subtle analysis of prescribing outliers. We found that STAR-PU likely underestimates prescribing to older adults, especially males and that a metric split by antibiotic family would better identify outliers. This analysis strengthens the case for updated STAR-PUs ^49^.

England is exceeding the WHO “60% Access” targets for antibiotic use, and likely has been since 2018, like other European settings^50^. However, this data shows the utility of knowing the variation by detailed age bands and sex: males aged 11-20 had less than 60% Access in 2015, rising to 70% in 2023 due to an increase in prescribing of the watch antibiotic lymecycline, likely for acne^36^. Whilst also spiking in females, due to the higher overall prescribing in women, their proportion Access remains above 70%.

Interventions to reduce antibiotic prescribing, including the Quality Premium, appear to be working in general in England, with most antibiotics (61%) having at least a 10% reduction in prescriptions between 2019 and 2023 ^51,52^. Disaggregating by age and sex reveals subgroups where changes in prescribing practice could be further targeted e.g. ofloxacin use has increased in men since 2016, potentially linked to rises in gonorrhoea^53^. Interestingly, a large reduction in prescriptions is not evidently driven by more ages being eligible for influenza vaccination: despite roughly 50% vaccine coverage we saw little prescription reduction in that ‘flu season in the age group vaccinated nor in linked age groups known to benefit from ‘flu vaccination of children (e.g. 65+ age band) ^54,55^.

These results are lower than those previously reported (e.g. 29% reduction in antibiotic prescribing in one systematic review ^22^) potentially due to our grouping antibiotics together, using wider age bands (to match vaccine targets) and not considering influenzas complexity (e.g. strain variation). The impact of influenza vaccinations may also be higher in other non-UK settings.

When considering AMR selection, we found no obvious link between the age and sex disaggregated exposures of beta-lactam antibiotics in primary care and higher MRSA prevalence in bloodstream infections (BSIs) in England. However, MRSA is often a hospital-associated pathogen ^56^, and BSIs are usually treated in hospitals, hence a lack of a strong link might have been expected but not no link. More broadly, we clearly see that women receive more antibiotics in primary care, which is at odds with higher male resistance rates in our previous research in BSIs across Europe ^25^ and available UK data^33^. Such patterns point to unexplained complexity in the pathways to resistance acquisition and potential intervention targets such as increased age or even sex-based prescribing recommendations. Untangling the link from ABU to AMR will require further age and sex disaggregated data but also information on other covariables such as patient ethnicity, deprivation level and co-morbidities ^18,57^.

Whilst the data used here is unparalleled in its detail and completeness, more patient data was available at later time points (a total of 3.3% of prescriptions were excluded), leading us to focus on the more complete 2023 data and we had to use sensitivity analyses to explore the hidden low numbers of prescriptions. Our data improves upon previous studies which have banded either patient age (in 10+ age bands), not considered disaggregation by sex or only focused on a subset of patient diagnoses ^7–9^. Moreover, much work in the UK is not all national data, using either only a subset of GPs ^8,9,57,58^ which although designed to be representative, may miss subtle geographical variation or incomplete market coverage (e.g. IQVIA has 89% market coverage in the UK ^50^). However, here we only had access to prescription data, with no measure of the propensity to take the prescribed medicine and whether this varies by age, sex, time or drug, no linked diagnosis or reason for prescribing, and only “Items”, rather than a more specific quantification, such as daily defined doses (DDDs) or grams ^59^. This is important as dose and duration of exposure is potentially relevant to the development of resistance ^60–62^.

The generalisability of this analysis to other locations is likely low (except in related geographies e.g. Wales, Scotland etc.), as the results may not be applicable to countries with different prescribing guidelines and healthcare systems (e.g. US insurance-based prescribing ^63^, or over-the-counter availability in LMICs ^64^). However, many of the underlying infection syndromes and behaviour patterns may exist and hence our findings add to the increasing evidence that we need disaggregated data by age and sex to optimally intervene against AMR.

Our analysis highlights the possibilities of using FOI requests in research. Future researchers could request additional UK data, e.g. at the GP level over larger time periods to avoid the issue of small numbers, as well as exploring requests for further (ideally linked) data that could help answer questions on health inequality such as socio-economic status and secondary health care infrastructure ^65^.

Overall, by showing wide antibiotic prescription variation across age and sex this research has demonstrated the need for a nuanced approach when exploring and understanding trends in ABU. Understanding how this affects the global public health problem that is AMR can only be done with matched age and sex disaggregated AMR data otherwise the impact of ABU targets, ageing populations and future interventions will be very hard to determine.

## Data Availability

All data are available at

https://opendata.nhsbsa.net/dataset/foi-01671

https://opendata.nhsbsa.net/dataset/foi-01975

## Acknowledgments

We would like to thank Yasmin Gant for supporting our understanding of reasons for antibiotic prescriptions, and Stephanie Evans for providing feedback on the manuscript.

## Transparency declarations

No conflicts to declare.

