## Supplement for "Antibiotic prescribing patterns by age and sex in England: why we need to take this variation into account to evaluate antibiotic stewardship and AMR selection"

### 1. Drug classifications

Table 1: Drug classifications. Aware Categories from [1]

| **BNF chemical substance code** | **Chemnical substance BNF DE=escription** | **Drug type** | **AWaRe Classification** |
| --- | --- | --- | --- |
| 0501011J0 | Benzylpenicillin sodium (Penicillin G) | Penicillins | Access |
| 0501011P0 | Phenoxymethylpenicillin (Penicillin V) | Penicillins | Access |
| 0501012G0 | Flucloxacillin sodium | Penicillins | Access |
| 0501012H0 | Flucloxacillin magnesium | Penicillins | Access |
| 0501012U0 | Temocillin sodium | Penicillins | Watch |
| 0501013B0 | Amoxicillin | Penicillins | Access |
| 0501013C0 | Amoxicillin sodium | Penicillins | Access |
| 0501013E0 | Ampicillin | Penicillins | Access |
| 0501013K0 | Co-amoxiclav (Amoxicillin/clavulanic acid) | Penicillins | Access |
| 0501013L0 | Co-fluampicil(Flucloxacillin/ampicillin) | Penicillins | Access |
| 0501014N0 | Ticarcillin with clavulanic acid | Penicillins | Watch |
| 0501014S0 | Piperacillin sodium/tazobactam sodium | Penicillins | Watch |
| 0501015P0 | Pivmecillinam hydrochloride | Penicillins | Access |
| 0501021A0 | Cefaclor | Cephalosporins and other beta-lactams | Watch |
| 0501021B0 | Cefadroxil | Cephalosporins and other beta-lactams | Access |
| 0501021C0 | Cefixime | Cephalosporins and other beta-lactams | Watch |
| 0501021D0 | Cefotaxime sodium | Cephalosporins and other beta-lactams | Watch |
| 0501021E0 | Cefoxitin sodium | Cephalosporins and other beta-lactams | Watch |
| 0501021F0 | Cefpodoxime | Cephalosporins and other beta-lactams | Watch |
| 0501021G0 | Ceftriaxone sodium | Cephalosporins and other beta-lactams | Watch |
| 0501021H0 | Ceftazidime pentahydrate | Cephalosporins and other beta-lactams | Watch |
| 0501021J0 | Cefuroxime sodium | Cephalosporins and other beta-lactams | Watch |
| 0501021K0 | Cefuroxime axetil | Cephalosporins and other beta-lactams | Watch |
| 0501021L0 | Cefalexin | Cephalosporins and other beta-lactams | Access |
| 0501021M0 | Cefradine | Cephalosporins and other beta-lactams | Access |
| 0501022A0 | Meropenem | Cephalosporins and other beta-lactams | Watch |
| 0501022B0 | Ertapenem sodium | Cephalosporins and other beta-lactams | Watch |
| 0501022D0 | Imipenem with cilastatin | Cephalosporins and other beta-lactams | Watch |
| 0501023A0 | Aztreonam | Cephalosporins and other beta-lactams | Reserve |
| 0501030F0 | Demeclocycline hydrochloride | Tetracyclines | Watch |
| 0501030I0 | Doxycycline hyclate | Tetracyclines | Access |
| 0501030L0 | Lymecycline | Tetracyclines | Watch |
| 0501030P0 | Minocycline hydrochloride | Tetracyclines | Watch |
| 0501030T0 | Oxytetracycline | Tetracyclines | Watch |
| 0501030V0 | Tetracycline | Tetracyclines | Access |
| 0501030X0 | Tetracycline combined preparations | Tetracyclines | Access |
| 0501030Y0 | Tigecycline | Tetracyclines | Reserve |
| 0501030Z0 | Doxycycline monohydrate | Tetracyclines | Access |
| 0501040C0 | Amikacin | Aminoglycosides | Access |
| 0501040H0 | Gentamicin sulfate | Aminoglycosides | Access |
| 0501040N0 | Neomycin sulfate | Aminoglycosides | Watch |
| 0501040U0 | Tobramycin | Aminoglycosides | Watch |
| 0501050A0 | Azithromycin | Macrolides | Watch |
| 0501050B0 | Clarithromycin | Macrolides | Watch |
| 0501050C0 | Erythromycin | Macrolides | Watch |
| 0501050H0 | Erythromycin ethylsuccinate | Macrolides | Watch |
| 0501050K0 | Erythromycin lactobionate | Macrolides | Watch |
| 0501050N0 | Erythromycin stearate | Macrolides | Watch |
| 0501060D0 | Clindamycin hydrochloride | Clindamycin and lincomycin | Access |
| 0501060E0 | Clindamycin phosphate | Clindamycin and lincomycin | Access |
| 0501070AA | Taurolidine | Some other antibacterials | NA |
| 0501070AB | Nitazoxanide | Some other antibacterials | NA |
| 0501070AC | Fidaxomicin | Some other antibacterials | Watch |
| 0501070AD | Tedizolid | Some other antibacterials | Reserve |
| 0501070AE | Fosfomycin trometamol | Some other antibacterials | Watch |
| 0501070F0 | Chloramphenicol | Some other antibacterials | Access |
| 0501070H0 | Colistin sulfate | Some other antibacterials | Reserve |
| 0501070I0 | Colistimethate sodium | Some other antibacterials | Reserve |
| 0501070M0 | Fusidic acid | Some other antibacterials | Watch |
| 0501070N0 | Sodium fusidate | Some other antibacterials | Watch |
| 0501070T0 | Teicoplanin | Some other antibacterials | Watch |
| 0501070U0 | Vancomycin hydrochloride | Some other antibacterials | Watch |
| 0501070W0 | Linezolid | Some other antibacterials | Reserve |
| 0501070X0 | Rifaximin | Some other antibacterials | Watch |
| 0501070Y0 | Daptomycin | Some other antibacterials | Reserve |
| 0501070Z0 | Pristinamycin | Some other antibacterials | Watch |
| 0501080D0 | Co-trimoxazole(Trimethoprim/sulfamethoxazole) | Sulfonamides and trimethoprim | Access |
| 0501080J0 | Sulfadiazine | Sulfonamides and trimethoprim | Access |
| 0501080T0 | Sulfamethoxypyridazine | Sulfonamides and trimethoprim | Access |
| 0501080V0 | Sulfapyridine | Sulfonamides and trimethoprim | Access |
| 0501080W0 | Trimethoprim | Sulfonamides and trimethoprim | Access |
| 0501090A0 | Aminosalicylic acid | Antituberculosis drugs | NA |
| 0501090C0 | Capreomycin | Antituberculosis drugs | Access |
| 0501090E0 | Cycloserine | Antituberculosis drugs | NA |
| 0501090H0 | Ethambutol hydrochloride | Antituberculosis drugs | NA |
| 0501090K0 | Isoniazid | Antituberculosis drugs | NA |
| 0501090N0 | Pyrazinamide | Antituberculosis drugs | NA |
| 0501090Q0 | Rifabutin | Antituberculosis drugs | Watch |
| 0501090R0 | Rifampicin | Antituberculosis drugs | Watch |
| 0501090S0 | Rifampicin combined preparations | Antituberculosis drugs | Watch |
| 0501090U0 | Streptomycin | Antituberculosis drugs | Watch |
| 0501090V0 | Bedaquiline | Antituberculosis drugs | NA |
| 0501100C0 | Clofazimine | Antileprotic drugs | NA |
| 0501100H0 | Dapsone | Antileprotic drugs | NA |
| 0501100J0 | Thalidomide (Antileprotic) | Antileprotic drugs | NA |
| 0501110C0 | Metronidazole | Metronidazole, tinidazole and ornidazole | Access |
| 0501110G0 | Tinidazole | Metronidazole, tinidazole and ornidazole | Access |
| 0501120L0 | Ciprofloxacin | Quinolones | Watch |
| 0501120N0 | Nalidixic acid | Quinolones | NA |
| 0501120P0 | Ofloxacin | Quinolones | Watch |
| 0501120Q0 | Norfloxacin | Quinolones | Watch |
| 0501120X0 | Levofloxacin | Quinolones | Watch |
| 0501120Y0 | Moxifloxacin | Quinolones | Watch |
| 0501130H0 | Methenamine hippurate | Urinary-tract infections | NA |
| 0501130R0 | Nitrofurantoin | Urinary-tract infections | Access |
| 0501130S0 | Fosfomycin calcium | Urinary-tract infections | Watch |

### 3. Unknowns

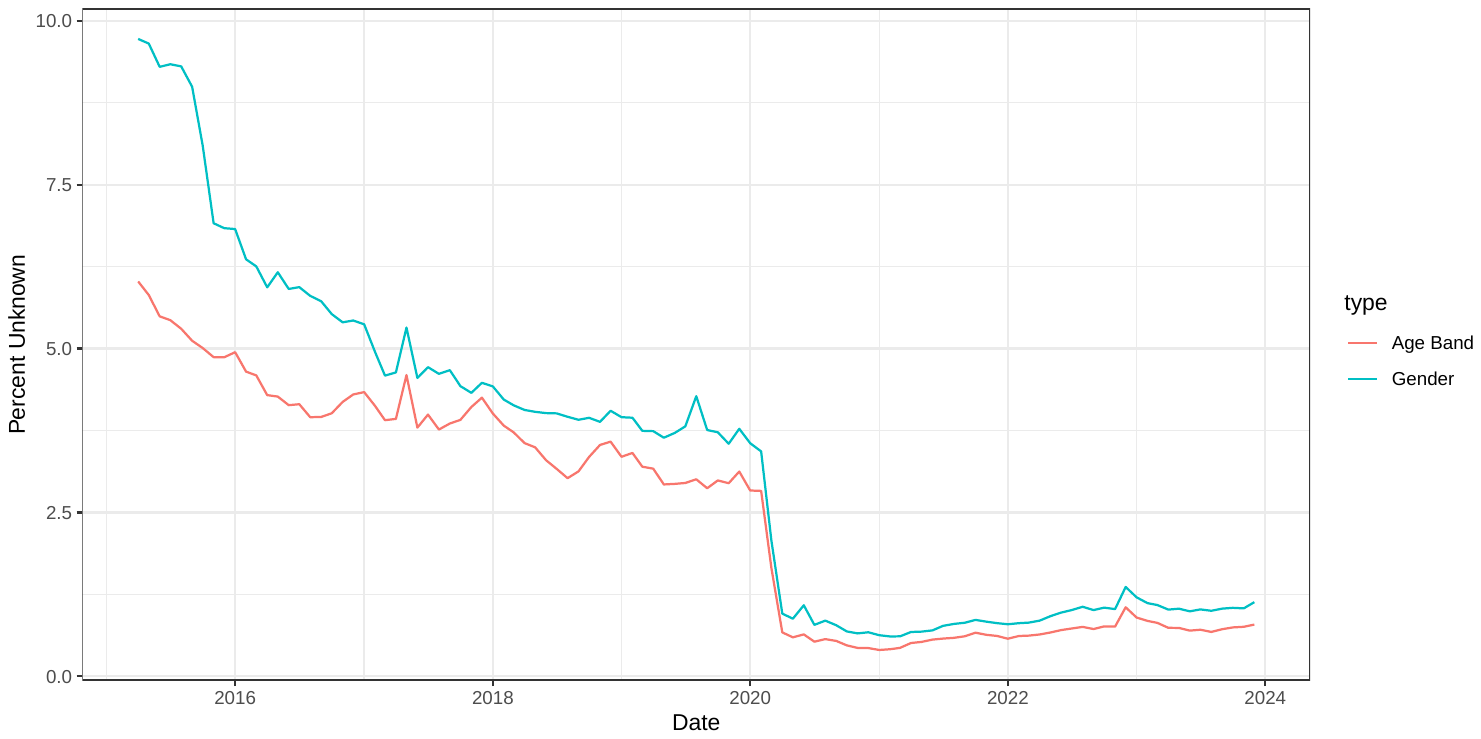

Figure 1: Percentage of total prescriptions that have "Unknown" Age band or Gender over time

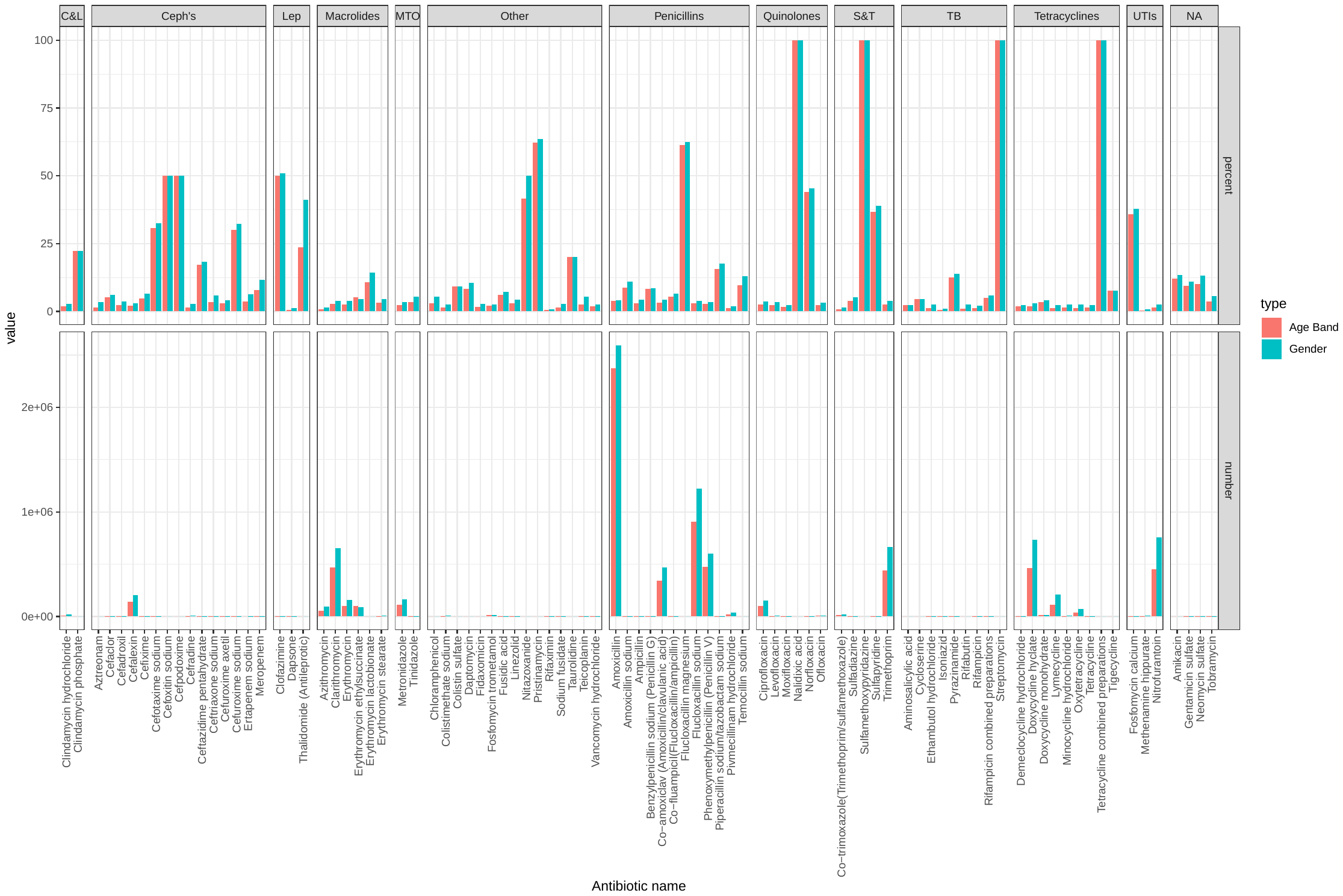

Figure 2: Percentage and Number of Prescription Items with Age Band or Gender as "Unknown"

### 5. Excluded antibiotics

The following antibiotics were excluded from the analysis, as they contained less than an average of 10 prescriptions per year (90 in total): temocillin sodium, tigecycline, colistin sulfate, daptomycin, pristinamycin, nitazoxanide, cycloserine, cefoxitin sodium, clindamycin phosphate, taurolidine, erythromycin lactobionate, capreomycin, thalidomide (antileprotic), imipenem with cilastatin, cefpodoxime, streptomycin, tetracycline combined preparations, nalidixic acid, ticarcillin with clavulanic acid, tedizolid, sulfamethoxypyridazine and bedaquiline.

### 6. Population data

Population sizes for England by age (up to age band 90+) and sex were accessed from the Office for National Statistics (ONS)[2], and the mid-year estimates applied to the prescription values for each calendar year, to calculate rates of prescription. ICB populations by age and sex were obtained from the ONS, using mid-year 2022 estimates[3] for England and only prescriptions for 2023 were used for sub-national analyses, due to changes in the definitions of the ICBs. Note that data from NHSBSA provides “Gender”, but most other sources use “Sex”. We have assumed that we can match these two in this analysis.

### 7. Combined prescriptions

Figure 3 shows the prescription rate per 100’000 population across all antibiotics. The 86+ age group has the highest prescription in both sexes, and the rate is higher in females than in males.

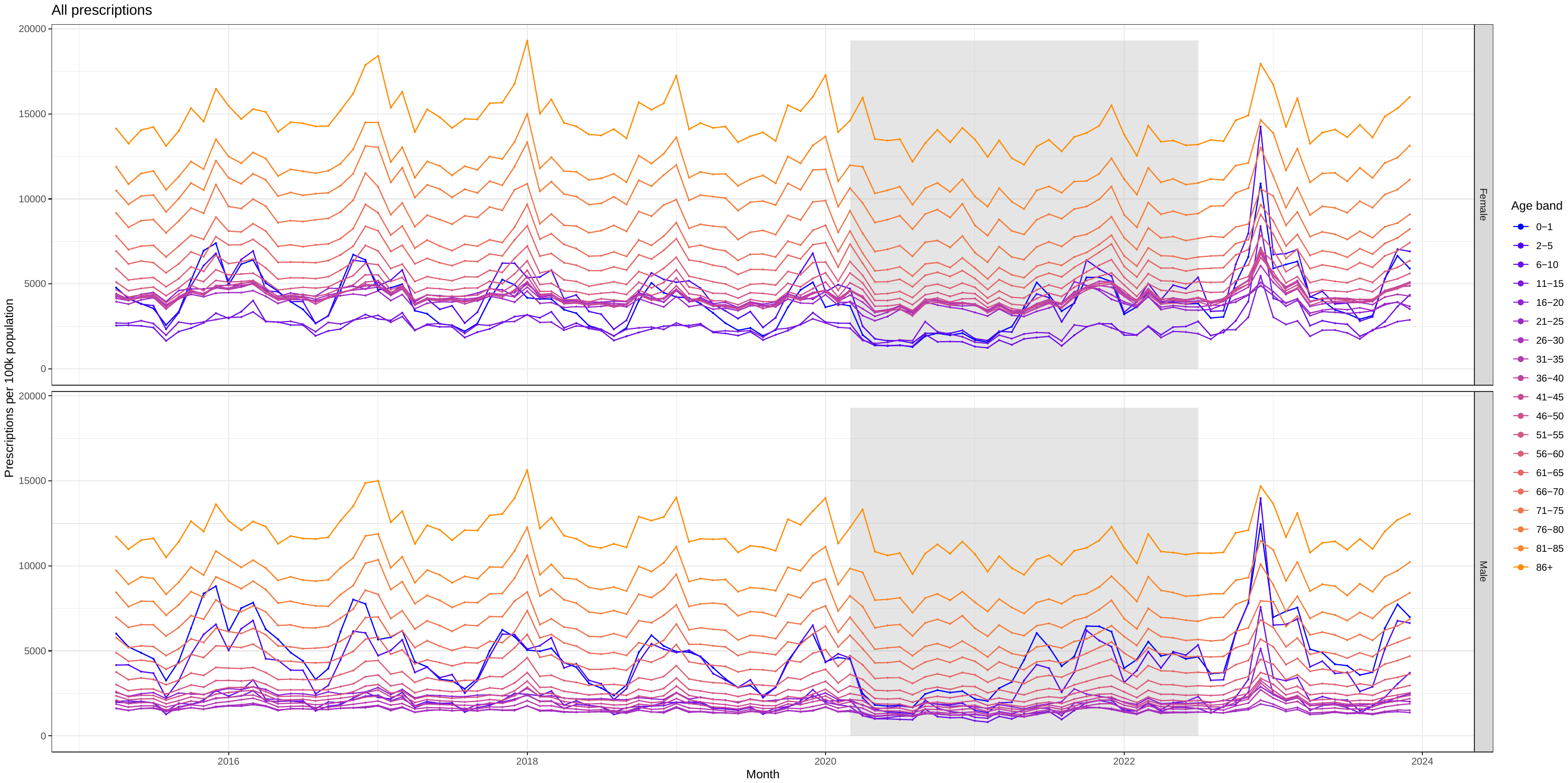

Figure 3: Prescription rate per 100’000 population across all antibiotic prescriptions. Colours indicate age groups, facets indicate sex. Grey shading indicates years of Covid-19 interventions.

### 8. Drugs overview

Table 2: Summary data on number of prescriptions for all prescribed antibiotics across full years in the data (2016-2023) unless otherwise indicated. Red shading indicates more prescriptions are to females. Top month is the month with the most prescriptions of this antibiotic when summing across the whole dataset (1 = January). Top age group (2023) is the age group in 2023 with the highest prescription rate per 100,000. Light green shading indicates relatively more prescriptions to younger individuals vs top age groups all being 55yo or more.

| Antibiotic | Total prescriptions | Mean annual prescriptions | % to females | Month | 2016 | 2019 | 2023 | Females | Males |
| --- | --- | --- | --- | --- | --- | --- | --- | --- | --- |
| Amoxicillin | 53,415,335 | 6,676,917 | 58 | 12 | 8,130,012 | 6,673,263 | 7,311,226 | 0-1,2-5,86+ | 0-1,2-5,86+ |
| Nitrofurantoin | 27,953,870 | 3,494,234 | 83 | 10 | 2,358,297 | 3,740,424 | 3,808,622 | 76-80,81-85,86+ | 76-80,81-85,86+ |
| Flucloxacillin sodium | 26,940,364 | 3,367,546 | 55 | 7 | 3,488,057 | 3,270,904 | 3,384,802 | 76-80,81-85,86+ | 76-80,81-85,86+ |
| Doxycycline hyclate | 22,868,632 | 2,858,579 | 60 | 12 | 2,415,657 | 2,772,514 | 3,804,564 | 76-80,81-85,86+ | 76-80,81-85,86+ |
| Phenoxymethylpenicillin (Penicillin V) | 15,557,160 | 1,944,645 | 59 | 12 | 1,892,172 | 1,841,679 | 2,447,555 | 16-20,2-5,6-10 | 0-1,2-5,6-10 |
| Trimethoprim | 14,439,590 | 1,804,949 | 75 | 1 | 3,087,875 | 1,488,922 | 1,428,569 | 76-80,81-85,86+ | 76-80,81-85,86+ |
| Clarithromycin | 14,417,677 | 1,802,210 | 63 | 1 | 2,077,261 | 1,845,315 | 1,743,846 | 76-80,81-85,86+ | 76-80,81-85,86+ |
| Co-amoxiclav (Amoxicillin/clavulanic acid) | 9,181,476 | 1,147,684 | 60 | 1 | 1,304,696 | 1,094,885 | 1,092,620 | 76-80,81-85,86+ | 76-80,81-85,86+ |
| Lymecycline | 7,938,074 | 992,259 | 58 | 3 | 1,054,631 | 1,032,182 | 861,423 | 11-15,16-20,21-25 | 11-15,16-20,21-25 |
| Cefalexin | 5,910,292 | 738,786 | 80 | 12 | 748,696 | 668,339 | 816,579 | 76-80,81-85,86+ | 76-80,81-85,86+ |
| Azithromycin | 5,877,890 | 734,736 | 57 | 12 | 612,414 | 748,885 | 805,700 | 71-75,76-80,81-85 | 76-80,81-85,86+ |
| Metronidazole | 4,156,192 | 519,524 | 78 | 3 | 599,382 | 520,332 | 474,629 | 21-25,26-30,31-35 | 76-80,81-85,86+ |
| Ciprofloxacin | 3,564,433 | 445,554 | 48 | 1 | 532,816 | 445,243 | 355,804 | 76-80,81-85,86+ | 76-80,81-85,86+ |
| Erythromycin | 3,397,046 | 424,631 | 66 | 1 | 709,522 | 394,861 | 286,163 | 76-80,81-85,86+ | 76-80,81-85,86+ |
| Oxytetracycline | 2,432,724 | 304,090 | 51 | 3 | 463,514 | 310,851 | 178,452 | 61-65,66-70,76-80 | 71-75,76-80,81-85 |
| Pivmecillinam hydrochloride | 1,790,378 | 223,797 | 79 | 10 | 108,361 | 223,030 | 301,447 | 76-80,81-85,86+ | 76-80,81-85,86+ |
| Erythromycin ethylsuccinate | 1,576,325 | 197,041 | 50 | 12 | 341,227 | 171,870 | 164,552 | 0-1,2-5,6-10 | 0-1,2-5,6-10 |
| Co-trimoxazole(Trimethoprim/sulfamethoxazole) | 1,417,031 | 177,129 | 44 | 12 | 129,051 | 173,722 | 228,051 | 66-70,76-80,81-85 | 76-80,81-85,86+ |
| Methenamine hippurate | 1,060,060 | 132,508 | 80 | 12 | 40,789 | 82,081 | 319,013 | 76-80,81-85,86+ | 76-80,81-85,86+ |
| Rifaximin | 593,273 | 74,159 | 41 | 12 | 33,589 | 71,036 | 109,205 | 56-60,61-65,66-70 | 56-60,61-65,66-70 |
| Clindamycin hydrochloride | 559,407 | 69,926 | 58 | 7 | 65,168 | 73,550 | 63,157 | 76-80,81-85,86+ | 76-80,81-85,86+ |
| Fosfomycin trometamol | 519,166 | 64,896 | 81 | 12 | 1,526 | 50,374 | 139,929 | 76-80,81-85,86+ | 76-80,81-85,86+ |
| Doxycycline monohydrate | 322,977 | 40,372 | 61 | 12 | 36,498 | 37,681 | 50,014 | 76-80,81-85,86+ | 76-80,81-85,86+ |
| Colistimethate sodium | 272,903 | 34,113 | 57 | 3 | 38,110 | 34,340 | 30,210 | 71-75,76-80,81-85 | 71-75,76-80,81-85 |
| Ofloxacin | 267,011 | 33,376 | 35 | 11 | 24,043 | 30,518 | 38,892 | 21-25,26-30,31-35 | 36-40,41-45,56-60 |
| Dapsone | 201,915 | 25,239 | 46 | 3 | 27,864 | 25,739 | 22,091 | 76-80,81-85,86+ | 76-80,81-85,86+ |
| Minocycline hydrochloride | 197,401 | 24,675 | 52 | 1 | 43,137 | 25,103 | 11,906 | 56-60,61-65,66-70 | 66-70,76-80,86+ |
| Tetracycline | 190,483 | 23,810 | 54 | 3 | 32,609 | 23,046 | 18,459 | 61-65,66-70,76-80 | 71-75,76-80,81-85 |
| Cefradine | 184,349 | 23,044 | 79 | 1 | 40,424 | 21,429 | 13,170 | 76-80,81-85,86+ | 76-80,81-85,86+ |
| Levofloxacin | 171,821 | 21,478 | 55 | 1 | 16,050 | 22,211 | 27,213 | 76-80,81-85,86+ | 76-80,81-85,86+ |
| Rifampicin | 128,935 | 16,117 | 54 | 1 | 20,217 | 16,708 | 12,220 | 76-80,81-85,86+ | 76-80,81-85,86+ |
| Isoniazid | 105,332 | 13,166 | 37 | 1 | 19,284 | 17,599 | 3,030 | 51-55,61-65,66-70 | 56-60,61-65,66-70 |
| Erythromycin stearate | 94,360 | 11,795 | 63 | 1 | 24,488 | 13,688 | 4,363 | 56-60,66-70,81-85 | 16-20,61-65,86+ |
| Cefaclor | 57,032 | 7,129 | 74 | 1 | 13,401 | 7,529 | 3,359 | 76-80,81-85,86+ | 2-5,76-80,81-85 |
| Co-fluampicil(Flucloxacillin/ampicillin) | 52,355 | 6,544 | 51 | 7 | 19,583 | 5,320 | 13 | 41-45,71-75,86+ | 76-80,81-85,86+ |
| Demeclocycline hydrochloride | 42,697 | 5,337 | 56 | 10 | 4,053 | 5,732 | 4,960 | 76-80,81-85,86+ | 76-80,81-85,86+ |
| Vancomycin hydrochloride | 39,368 | 4,921 | 64 | 8 | 3,511 | 3,664 | 7,638 | 76-80,81-85,86+ | 76-80,81-85,86+ |
| Ethambutol hydrochloride | 35,446 | 4,431 | 53 | 1 | 6,694 | 5,166 | 1,872 | 71-75,76-80,81-85 | 61-65,71-75,86+ |
| Moxifloxacin | 32,559 | 4,070 | 56 | 1 | 4,842 | 4,721 | 2,919 | 61-65,76-80,81-85 | 61-65,76-80,81-85 |
| Cefuroxime axetil | 29,432 | 3,679 | 70 | 3 | 5,348 | 4,046 | 1,862 | 71-75,76-80,81-85 | 71-75,76-80,86+ |
| Gentamicin sulfate | 24,155 | 3,019 | 50 | 1 | 4,189 | 3,116 | 2,285 | 71-75,81-85,86+ | 76-80,81-85,86+ |
| Sodium fusidate | 15,454 | 1,932 | 39 | 3 | 3,338 | 1,973 | 790 | 71-75,81-85,86+ | 66-70,81-85,86+ |
| Cefadroxil | 14,884 | 1,860 | 85 | 3 | 5,313 | 822 | 242 | 71-75,81-85,86+ | 76-80,81-85,86+ |
| Cefixime | 8,149 | 1,019 | 61 | 1 | 2,004 | 918 | 505 | 21-25,56-60,76-80 | 71-75,81-85,86+ |
| Rifampicin combined preparations | 7,791 | 974 | 45 | 8 | 1,472 | 962 | 1,478 | 16-20,21-25,26-30 | 26-30,31-35,86+ |
| Linezolid | 6,723 | 840 | 54 | 6 | 555 | 831 | 1,031 | 76-80,81-85,86+ | 71-75,81-85,86+ |
| Tobramycin | 6,190 | 774 | 51 | 1 | 1,444 | 797 | 417 | 71-75,76-80,86+ | 71-75,76-80,81-85 |
| Ampicillin | 6,100 | 762 | 66 | 3 | 1,673 | 888 | 161 | 0-1,61-65,86+ | 0-1,81-85,86+ |
| Ceftriaxone sodium | 5,813 | 727 | 55 | 1 | 1,380 | 572 | 410 | 16-20,21-25,26-30 | 71-75,81-85,86+ |
| Tinidazole | 5,578 | 697 | 66 | 1 | 1,536 | 1,179 | 2 | NA | 31-35,36-40 |
| Teicoplanin | 4,928 | 616 | 56 | 6 | 1,237 | 486 | 216 | 76-80,81-85,86+ | 71-75,81-85,86+ |
| Benzylpenicillin sodium (Penicillin G) | 2,477 | 310 | 45 | 3 | 603 | 329 | 187 | 0-1,81-85,86+ | 0-1,81-85,86+ |
| Fidaxomicin | 1,979 | 247 | 64 | 9 | 98 | 103 | 656 | 76-80,81-85,86+ | 76-80,81-85,86+ |
| Sulfadiazine | 1,893 | 237 | 57 | 1 | 541 | 258 | 90 | 0-1,41-45,86+ | 0-1,71-75,86+ |
| Neomycin sulfate | 1,614 | 202 | 45 | 5 | 442 | 286 | 1 | 61-65 | NA |
| Rifabutin | 1,288 | 161 | 49 | 5 | 240 | 130 | 117 | 76-80,81-85,86+ | 66-70,76-80,81-85 |
| Fusidic acid | 1,116 | 140 | 46 | 6 | 210 | 156 | 66 | 0-1,81-85,86+ | 0-1,81-85,86+ |
| Meropenem | 995 | 124 | 49 | 3 | 202 | 123 | 77 | 76-80,81-85,86+ | 76-80,81-85,86+ |
| Ertapenem sodium | 810 | 101 | 59 | 4 | 178 | 71 | 40 | 76-80,81-85,86+ | 71-75,76-80,81-85 |
| Amoxicillin sodium | 782 | 98 | 48 | 1 | 153 | 99 | 51 | 0-1,66-70,86+ | 76-80,81-85,86+ |
| Pyrazinamide | 690 | 86 | 41 | 12 | 88 | 141 | 54 | 66-70,76-80,86+ | 51-55,66-70,71-75 |
| Chloramphenicol | 452 | 56 | 51 | 3 | 97 | 58 | 30 | 21-25,76-80,81-85 | 66-70,71-75,76-80 |
| Ceftazidime pentahydrate | 449 | 56 | 61 | 1 | 65 | 55 | 41 | 76-80,81-85,86+ | 61-65,71-75,86+ |
| Piperacillin sodium/tazobactam sodium | 271 | 34 | 67 | 1 | 52 | 35 | 15 | 26-30,71-75,81-85 | 86+ |
| Fosfomycin calcium | 242 | 30 | 67 | 7 | 63 | 35 | 10 | 41-45,71-75,76-80 | 66-70 |
| Cefotaxime sodium | 225 | 28 | 55 | 1 | 63 | 34 | 12 | 2-5,66-70,81-85 | 0-1,31-35,76-80 |
| Cefuroxime sodium | 219 | 27 | 85 | 8 | 38 | 23 | 43 | 21-25,36-40,41-45 | 51-55,56-60 |
| Aztreonam | 176 | 22 | 48 | 3 | 38 | 26 | 7 | 26-30 | 26-30 |
| Clofazimine | 167 | 21 | 41 | 6,7 | 18 | 39 | 21 | 16-20,56-60,76-80 | 51-55,61-65,76-80 |
| Amikacin | 156 | 20 | 49 | 3 | 92 | 6 | 9 | 26-30,46-50,66-70 | 61-65 |
| Aminosalicylic acid | 126 | 16 | 51 | 6 | 5 | 15 | 23 | 0-1,71-75,86+ | 61-65,71-75,76-80 |
| Norfloxacin | 112 | 14 | 79 | 7 | 14 | 37 | 1 | 51-55 | NA |
| Sulfapyridine | 92 | 12 | 32 | 5 | 25 | 10 | 17 | 86+ | 86+ |
| Flucloxacillin magnesium | 27 | 4 | 22 | 9 | 8 | 4 | 2 | 86+ | 2-5 |

### 10. Sensitivity Analyses *’s

In total four different scenarios were run with the use of *s (where the stars are hidden values between 1 and 4).

- Default scenario: all * values converted to 1s
- Sensitivity “four”: all * values converted to 4s
- Gender split: * for prescriptions to males converted to 4s, *s for prescriptions to females converted to 1s.
- Age split: *s for prescriptions to those aged 20 and under converted to 4s, *s for prescriptions to those over 20 converted to 1s.

The full analysis (all paper and appendix plots) for each is available on github. The figure 1 from the main paper for each scenario is produced below. All scenarios resulted in the same trends, with the different scenarios showing the trends to varying extents.

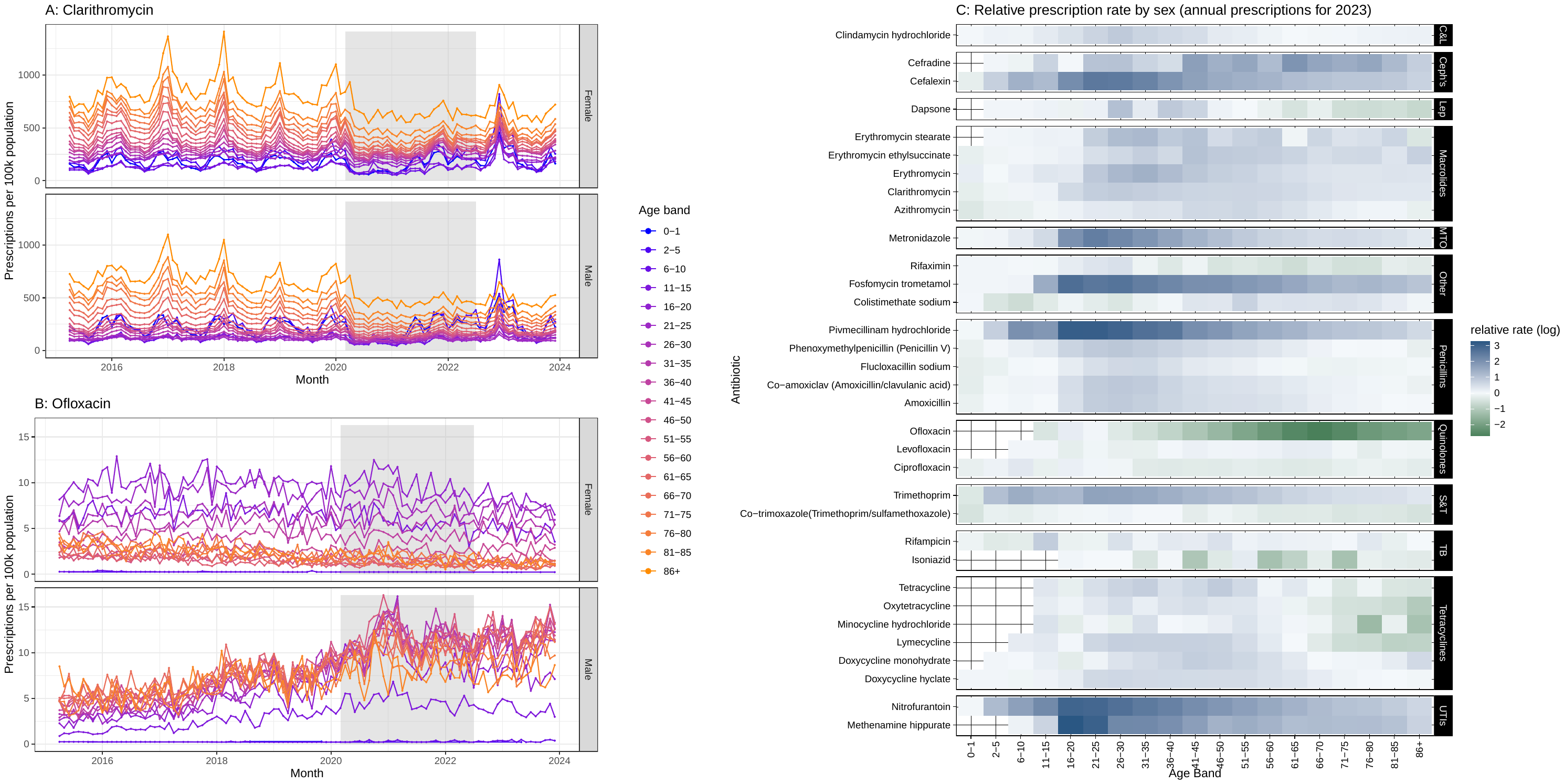

Figure 4: Sensitivity "four". A/B). Prescription rate per 100’000 population for Clarithromycin and Ofloxacin respectively. Colours indicate age groups, facets indicate sex. Grey shading indicates years of Covid-19 interventions. C) Relative prescription rate by sex for prescribed medicines in 2023. Colours represents the log of the ratio, with blue indicating higher prescriptions in women, and green indicating higher prescriptions in women. Only drugs with more than 100’000 prescriptions across the whole time period where included in the plot. Drugs are ordered by BNF classification, where the acronyms are: Cephalosporins and other beta-lactams = “Ceph's”, Clindamycin and lincomycin = "C&L", Sulphonamides and trimethoprim= “S&T”, Antileprotic drugs = “Lep”, Antituberculosis drugs = "TB", Urinary-tract infections = "UTIs", Metronidazole, tinidazole and ornidazole = "MTO".

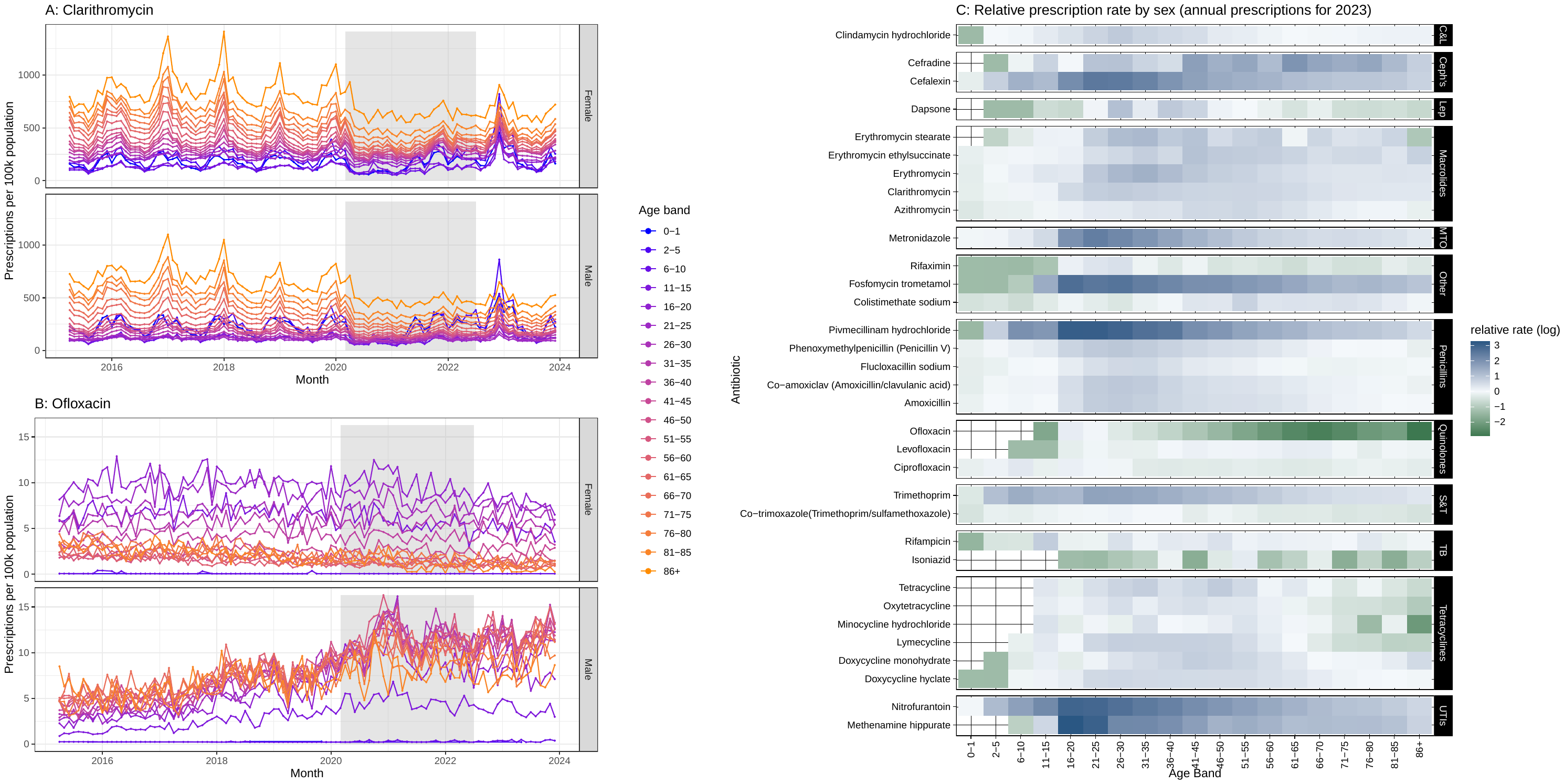

Figure 5: Gender split sensitivity. A/B). Prescription rate per 100’000 population for Clarithromycin and Ofloxacin respectively. Colours indicate age groups, facets indicate sex. Grey shading indicates years of Covid-19 interventions. C) Relative prescription rate by sex for prescribed medicines in 2023. Colours represents the log of the ratio, with blue indicating higher prescriptions in women, and green indicating higher prescriptions in women. Only drugs with more than 100’000 prescriptions across the whole time period where included in the plot. Drugs are ordered by BNF classification, where the acronyms are: Cephalosporins and other beta-lactams = “Ceph's”, Clindamycin and lincomycin = "C&L", Sulphonamides and trimethoprim= “S&T”, Antileprotic drugs = “Lep”, Antituberculosis drugs = "TB", Urinary-tract infections = "UTIs", Metronidazole, tinidazole and ornidazole = "MTO".

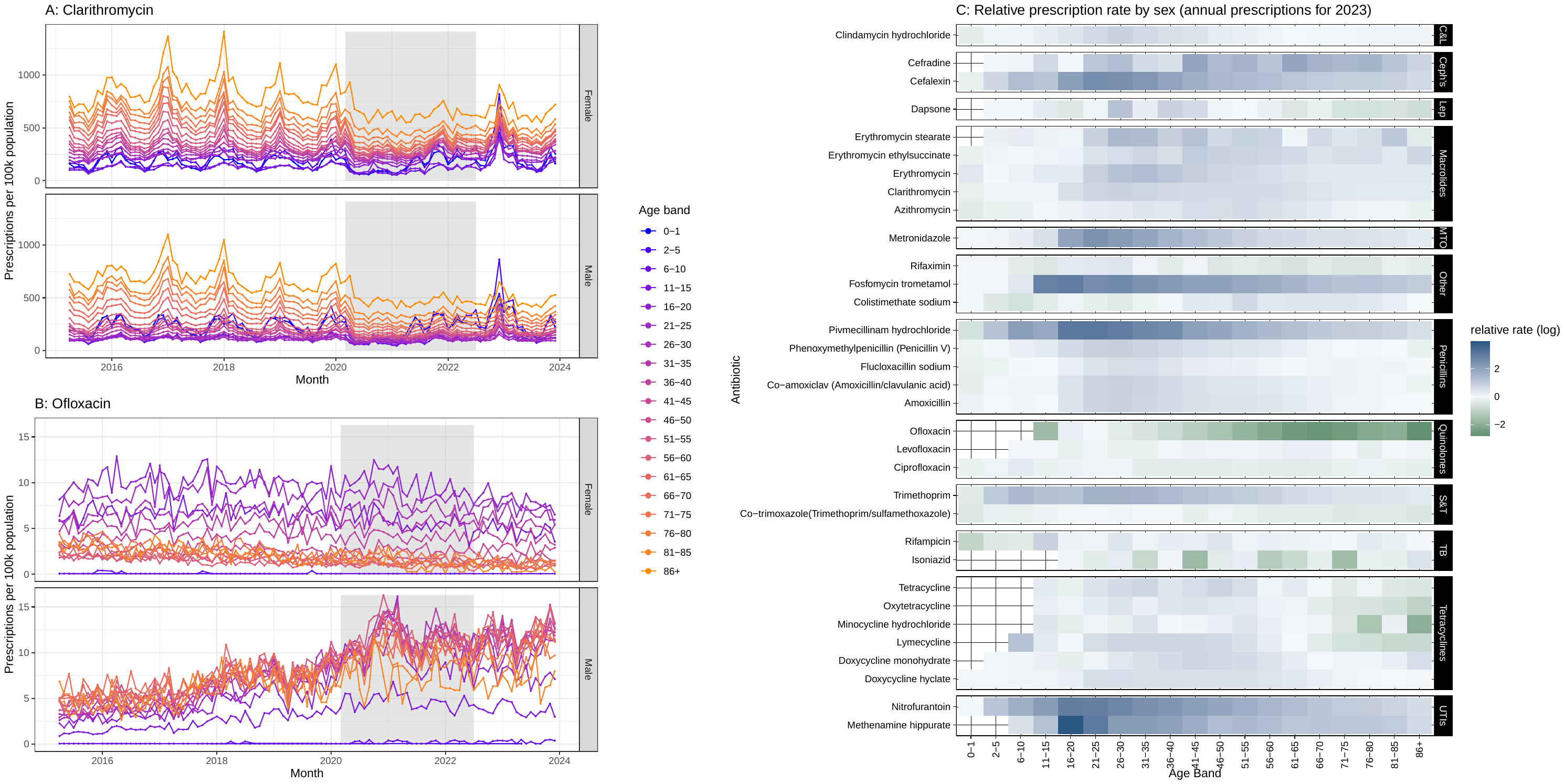

Figure 6: Age split sensitivity. A/B). Prescription rate per 100’000 population for Clarithromycin and Ofloxacin respectively. Colours indicate age groups, facets indicate sex. Grey shading indicates years of Covid-19 interventions. C) Relative prescription rate by sex for prescribed medicines in 2023. Colours represents the log of the ratio, with blue indicating higher prescriptions in women, and green indicating higher prescriptions in women. Only drugs with more than 100’000 prescriptions across the whole time period where included in the plot. Drugs are ordered by BNF classification, where the acronyms are: Cephalosporins and other beta-lactams = “Ceph's”, Clindamycin and lincomycin = "C&L", Sulphonamides and trimethoprim= “S&T”, Antileprotic drugs = “Lep”, Antituberculosis drugs = "TB", Urinary-tract infections = "UTIs", Metronidazole, tinidazole and ornidazole = "MTO".

### 11. UCM-family

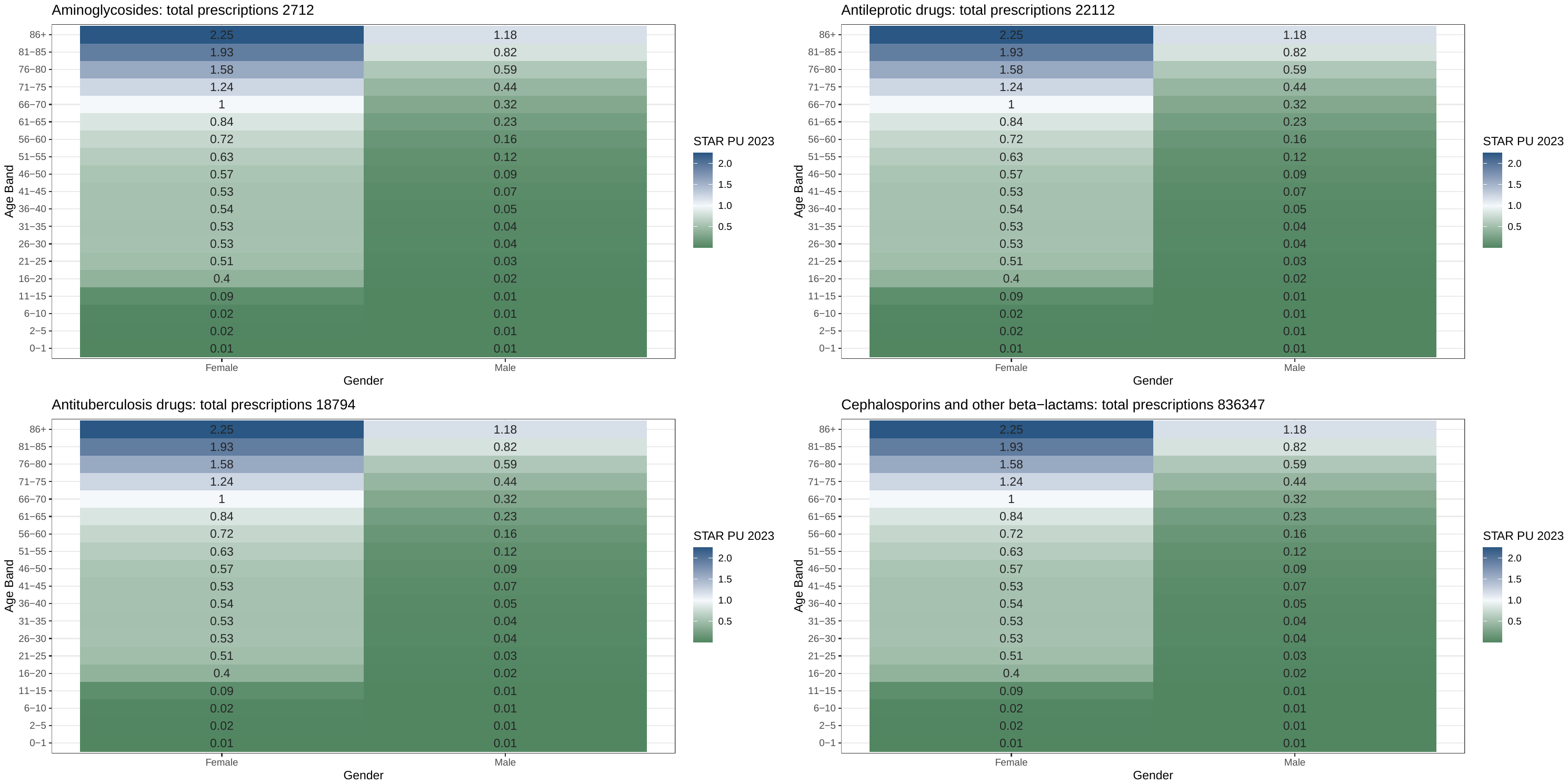

Figure 7: Updated Comparison Metric (UCM) for different drug families, using 2023 data and more disaggregated age bands. Blue indicates a high value, green a low value, centred around the base-age-sex band of females aged 66-70 (white)

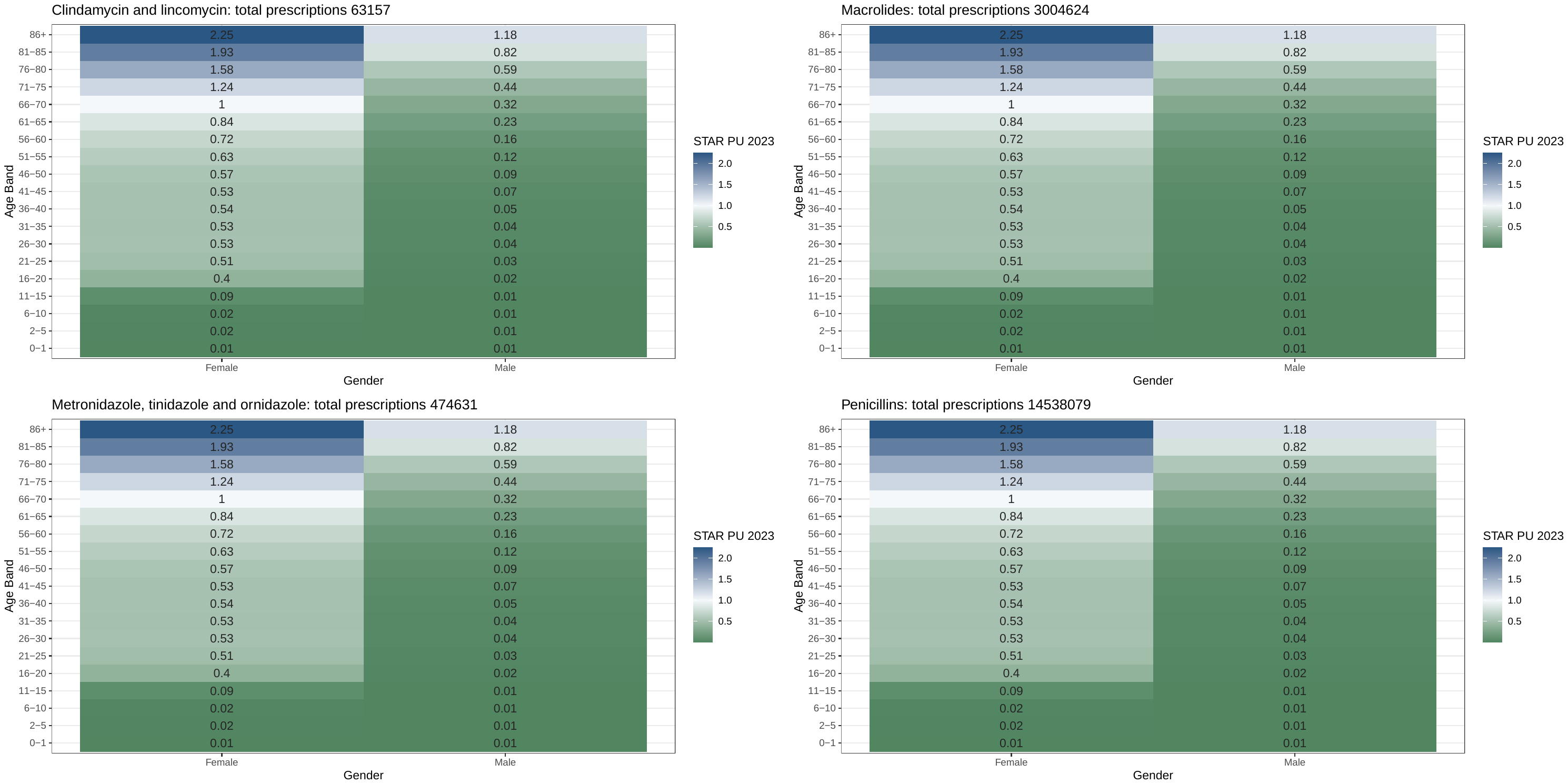

Figure 8: Updated Comparison Metric (UCM) for different drug families, using 2023 data and more disaggregated age bands. Blue indicates a high value, green a low value, centred around the base-age-sex band of females aged 66-70 (white)

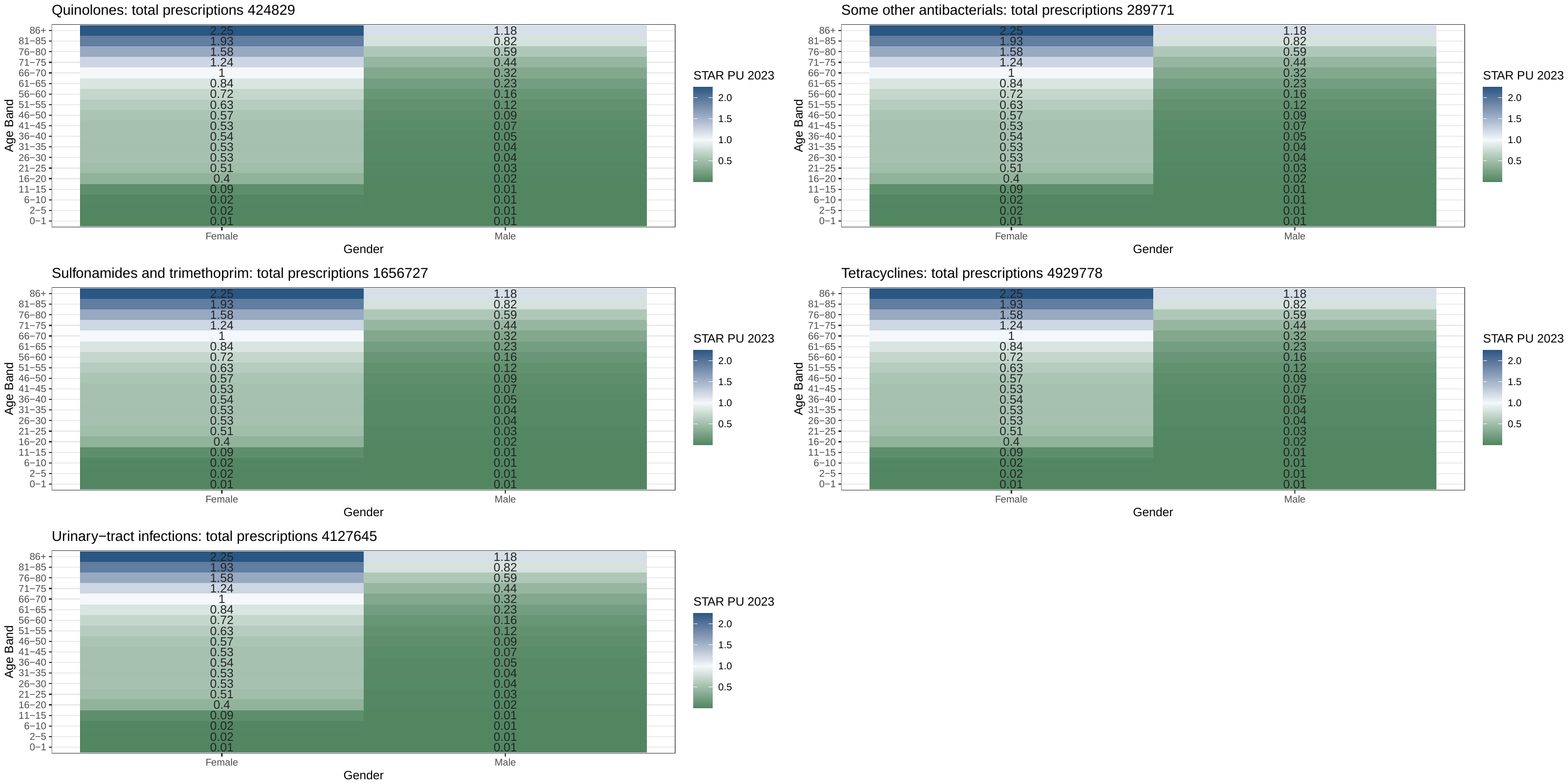

Figure 9: Updated Comparison Metric (UCM) for different drug families, using 2023 data and more disaggregated age bands. Blue indicates a high value, green a low value, centred around the base-age-sex band of females aged 66-70 (white)

### 12. UCM Ranking

Changes in rank order between the overall UCM and the drug family UCMs are displayed in Figure 6.

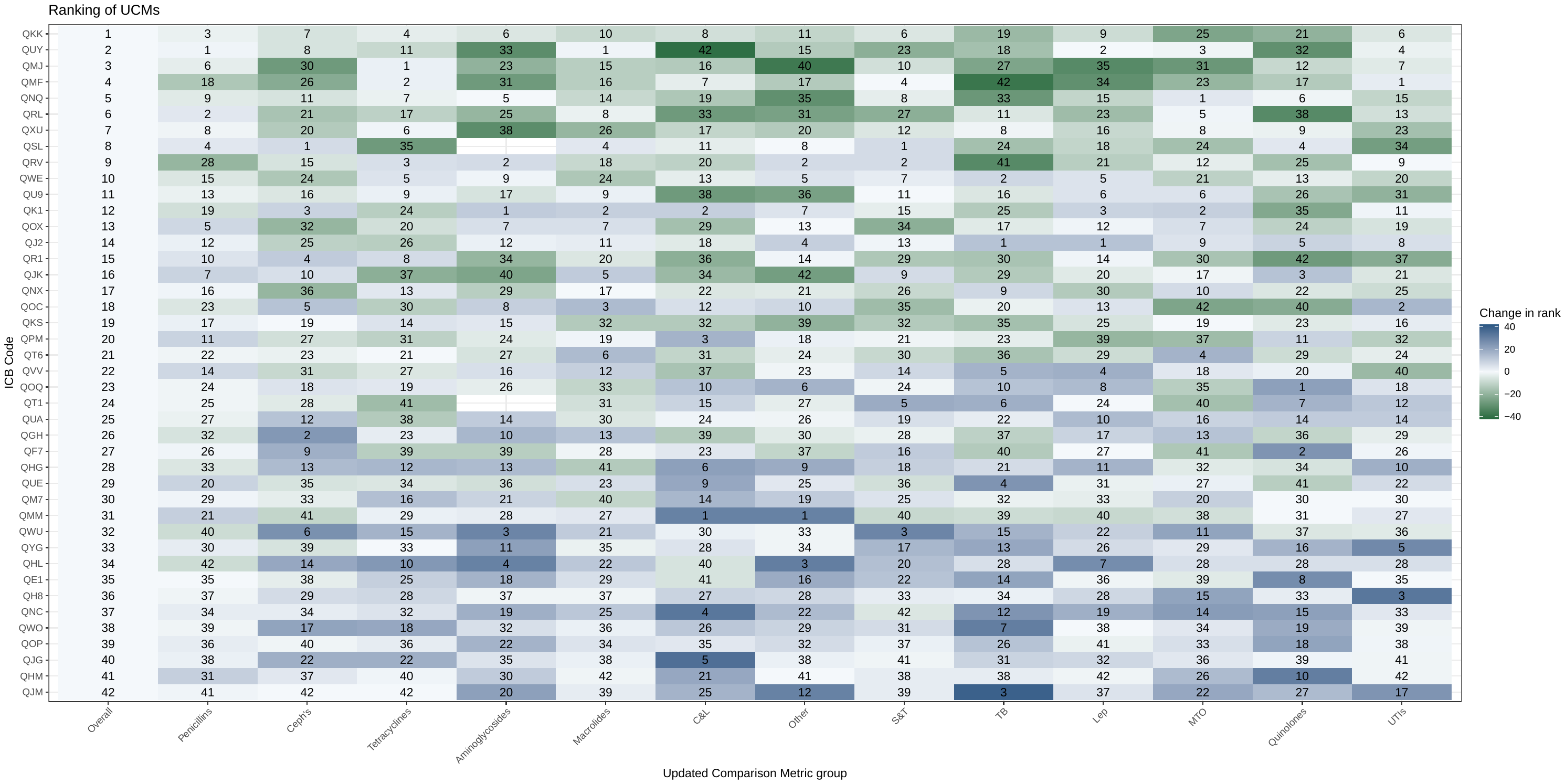

Figure 10: Ranking of ICMs. The “Overall” column shows the ranking of ICBs using our UCM, and the other columns show the ranking of ICBs by the drug family specific UCMs. Colour indicates the change in rank number from the overall ranking. Numbers indicate the rank. Acronyms: Cephalosporins and other beta-lactams = “Ceph's”, Clindamycin and lincomycin = "C&L", Sulfonamides and trimethoprim= “S&T”, Antileprotic drugs = “Lep”, Antituberculosis drugs = "TB", Urinary-tract infections = "UTIs", Metronidazole, tinidazole and ornidazole = "MTO".

### 13. AWaRe classifications by ICB

The percentage of prescriptions that were in the Access AWaRe class varied by ICB in 2023 (Figure 7). The difference in percentage Access prescribed varied across ICBs by between 5.3% (in females aged 6-10), to 13.4% (in males aged 16-20).

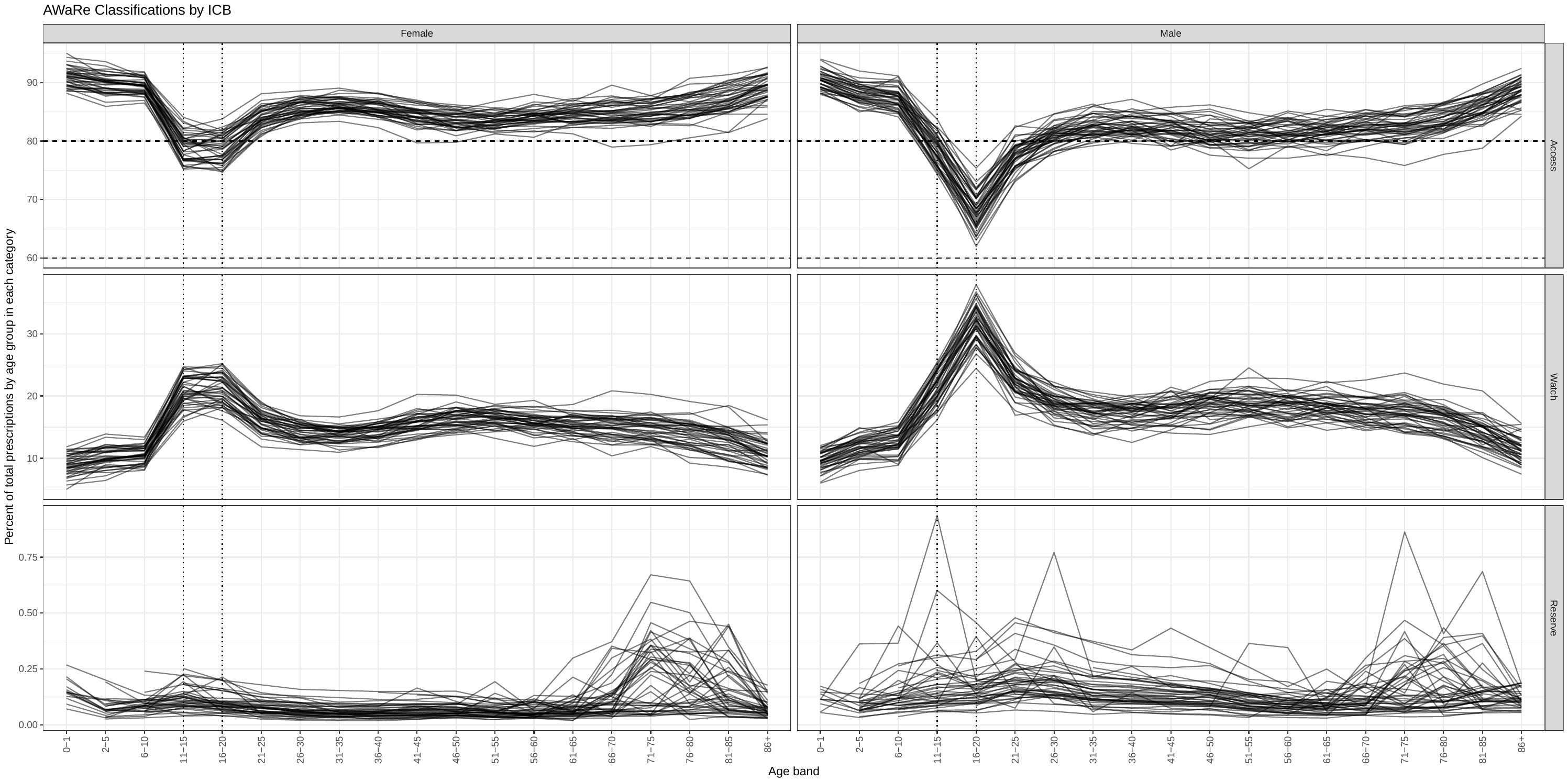

Figure 11: Percentage of total prescriptions in each of the Aware categories, by age group and sex in 2023. Lines indicate individual ICBs. Dashed horizontal lines represent targets of 60% and 80% in the Access category, and dotted vertical lines are included to aid comparison across graphs
